# Plasma proteomics reveals signatures of cognitive aging and response to the FINGER multidomain lifestyle intervention

**DOI:** 10.64898/2026.11.13.26367619

**Authors:** Makrina Daniilidou, Andrea L. Benedet, Ari Solomon, Anna Rosenberg, Ludivine Morvan, Jenni Lehtisalo, Francesca Mangialasche, Esko Levälahti, Burak Arslan, Markku Peltonen, Ilaria Pola, Kübra Tan, Tiia Ngandu, Henrik Zetterberg, Miia Kivipelto, Anna Matton

**Author notes:** Corresponding author: Dr. Anna Matton. These authors contributed equally (Henrik Zetterberg, Miia Kivipelto, Anna Matton).

## Abstract

The FINGER trial demonstrated that a multidomain lifestyle intervention benefits cognition in older adults at-risk of dementia, yet the underlying biological mechanisms remain unclear. We measured neurodegeneration-related proteins using the NULISAseq CNS disease panel in 1158 FINGER participants to investigate associations between circulating proteins, cognitive trajectories, and intervention response. Independent of the intervention, ten proteins spanning immune, neuronal, and metabolic processes were associated with poorer cognitive trajectories, with TREM2 among the strongest signals. The intervention led to changes in eight proteins, including CRH, IL-7 and MME. Eleven proteins modified the cognitive response to intervention, with KDR and CRH showing the strongest effect. Complementary Elastic Net analyses confirmed CRH, and further identified PTN and CCL2 as markers of intervention responsiveness. Additional proteins showed APOE ε4- and sex-specific associations with the intervention response. Together, these findings identify proteomic signatures of cognitive response that could inform precision dementia prevention.

## Introduction

The growing global burden of Alzheimer’s disease and related disorders (ADRD) represents a profound health challenge, underscoring the need for effective interventions for preserving cognitive function among people at risk of dementia. ADRDs are driven by complex disease mechanisms^1^, with modifiable lifestyle and health-related factors accounting for an estimated 45% of the overall risk^2^. Despite recent advances in disease-modifying therapies for early Alzheimer’s disease (AD), available treatments have modest clinical effects, are restricted to selected patient groups, and remain challenging to implement at population level ^3–5^. Thus, lifestyle-based interventions have emerged as a promising strategy to mitigate cognitive decline and potentially delay the onset of ADRD, as emphasized in the updated 2026 WHO guidelines on dementia risk reduction^6^. Dietary patterns, physical activity, cognitive and social engagement, and effective management of vascular and metabolic risk factors have all been linked to better cognitive health and reduced dementia risk^2,6,7^. The Finnish Geriatric Intervention Study to Prevent Cognitive Impairment and Disability (FINGER) was the first clinical trial which provided proof of concept that targeting these factors simultaneously can improve cognition and exert other related health benefits in older adults at increased risk of dementia^8,9^. This two-year landmark trial initiated the World-Wide FINGERS (WW-FINGERS) network of multidomain dementia risk reduction trials ^10^ throughout which the initial FINGER findings have been replicated in other geographical and cultural settings (e.g., in North America, US-POINTER^11^ and South America, Latam-FINGERS^12^). Yet an important question remains: what happens biologically when such an intervention has an impact on cognitive function, and why do some individuals benefit more than others?

Blood-based biomarkers provide a window into these processes. The rapid advances in the biomarker field offer important insights into cognitive health and present significant opportunities for understanding and further developing the early intervention strategies in the context of ADRD. Plasma phosphorylated tau, amyloid-β, glial fibrillary acidic protein and neurofilament light are biomarkers currently used for tracking neuropathological and neurodegenerative processes^13,14^. However, these established markers capture only a fraction of the molecular changes associated with cognitive aging and early neurodegenerative processes and may provide limited information on the broader biological mechanisms through which multidomain lifestyle interventions exert their effects^15^. Therefore, circulating proteins reflecting other molecular pathways that are central to brain health, such as inflammation, vascular function, metabolic homeostasis, and synaptic integrity, may help to understand how lifestyle interventions affect systemic and brain-related biology. They may also help identify individuals most likely to benefit from these interventions.

In this study, we characterized circulating protein profiles using the highly-sensitive, state-of-the-art NULISA technology^16^ (NULISAseq™ CNS Disease 120 panel) in the FINGER trial. The panel included both established ADRD biomarkers and a broader set of mostly less explored multi-mechanistic proteins. In a separate work we focused on the core markers^17^; here, we focused on proteins reflecting broader disease biology. We first investigated the associations of these proteins with cognitive performance and cognitive change in the whole population. We further evaluated intervention-related changes in protein levels over time, identified baseline proteins associated with intervention responsiveness and examined whether these associations differed according to *APOE ε4* carrier status and sex. Established ADRD biomarkers were examined separately as reference markers in correlation analyses. To capture the multidimensional nature of biological responses, we applied machine learning approaches to identify protein signatures predictive of intervention response. Finally, baseline protein profiles were compared between intervention-adherent responders and non-responders to identify molecular pathways associated with successful cognitive benefit from the lifestyle intervention.

## Methods

### Study Setting and Population

This research constitutes an exploratory analysis of the FINGER RCT (ClinicalTrials.gov ID NCT01041989), which evaluated the effectiveness of a multidomain lifestyle intervention on cognition among community-dwelling older adults at risk for dementia across six centers in Finland (N=1260, average age 69 years at baseline). Screening and baseline evaluations occurred between September 2009 and November 2011, and the core two-year RCT was completed in February 2014. Details on the FINGER protocol, and main trial findings are available elsewhere^8,9^. The study received ethical approval from the Coordinating Ethics Committee of Helsinki and Uusimaa Hospital District (94/13/03/00/09, HUS/1204/2017) and Swedish Ethical Review Authority (2020-07058), with all participants providing written informed consent.

Participants were recruited from prior national surveys, with inclusion criteria set at ages 60–77, a CAIDE Dementia Risk Score^18^ of at least six points, and cognitive function at or just below mean levels for their age. Key exclusion criteria included previously diagnosed or suspected dementia, significant cognitive impairment (Mini-Mental State Examination (MMSE) <20), and conditions that could compromise safe participation. This selection process aimed to represent the general Finnish older population with dementia risk factors but minimal cognitive impairment^19^.

Participants were randomly assigned in a 1:1 ratio to either the intervention group (intensive multidomain lifestyle program) or the control group (standard health advice). Randomization was executed using computer-generated allocations in blocks of four at each site (two participants to each group). Efforts were made to achieve double-blinding, with outcome assessors unaware of intervention specifics and group allocations remaining undisclosed. The primary outcome was change in global cognition measured by a total composite z-score from the Neuropsychological Test Battery (NTB). After two years, the intervention group exhibited greater cognitive improvement compared to controls and a reduced risk of cognitive decline.^9^

### Intervention

All participants received baseline lifestyle information (mini-intervention) and feedback regarding their vascular risk factors. Post-randomization, control participants were given routine health advice and risk factor information during main study visits at six months, one year, and two years, along with instructions to consult healthcare providers as needed. Those in the intervention group engaged in a comprehensive program encompassing dietary counseling, physical activity, cognitive stimulation, and monitoring of vascular and metabolic factors. The intervention details are described elsewhere^8,9^. Interventions were delivered by a multidisciplinary team in accordance with national and international guidelines. Intervention adherence was assessed according to a prespecified definition^20^ based on participation in the activities offered to the intervention group, with high adherence defined as >50% overall participation across intervention domains.

In summary, the dietary intervention included three individual sessions and six to eight group sessions tailored to participants’ needs. The exercise component included progressive strength training (one to three weekly group sessions) alongside individualized aerobic training (two to five weekly sessions). Cognitive training encompassed ten group sessions (educational content) and individualized computer-based activities (in two six-month blocks, training three times weekly), targeting executive function, memory, and processing speed with distinct tasks from outcome assessments. Management of metabolic and vascular risks sought to enhance blood pressure, lipids, glucose, and weight, adhering to national care guidelines. Six additional meetings provided further measurements and lifestyle guidance, with no medications prescribed; participants were advised to reach out to healthcare providers if necessary.

### Cognitive Assessment

Cognitive assessments using the NTB were carried out by blinded psychologists at baseline, one year, and two years. The trial’s primary outcome was the change in the NTB total score, representing global cognition (mean of 14 tests, with at least eight required for calculations). Scores at each timepoint were converted to z-scores standardized to baseline mean and SD (higher scores indicating better performance). Detailed description of the cognitive assessment has been published elsewhere^9^.

### Blood Sample Collection and Analysis

Fasting venous blood samples were obtained at baseline, six months, one year, and two years. Serum and plasma were separated immediately, frozen, and transferred monthly to the Finnish Institute for Health and Welfare laboratory, where samples were aliquoted and stored at −70°C for future analysis. APOE genotype was assessed using standard techniques^21^. Fluid biomarkers were analyzed in all available frozen plasma samples from baseline (n=1225) and the two-year follow-up (n=1105). Samples were shipped in February 2024 and analyzed at the Clinical Neurochemistry Laboratory at the University of

Gothenburg, Gothenburg, Sweden. The assays included the clinically validated Fujirebio Lumipulse test for p-tau217, and the NULISAseq™ CNS Disease 120 Panel (Alamar Biosciences). The NULISA methodology (Nucleic acid Linked Immuno-Sandwich Assay) allows ultrasensitive multiplex detection and quantification of approximately 120 proteins at attomolar concentrations^22^.

Participants samples were balanced (randomization group allocation and age) across study plates while keeping longitudinal samples of a single individual on the same plate. Sample were analyzed in the ARGO HT platform and resulting library pools sequenced on an Illumina NextSeq 2000 instrument (Illumina, Inc., San Diego, CA, USA), as specified by the manufacturer. The sequencing output was processed using Alamar’s proprietary software, which included rescaling and log transformation of the data, with the generation of NULISA Protein Quantification (NPQ) values ^23,24^. Therefore, NPQ units reflect relative protein abundances and differences are interpreted as log2 (fold change), whereas the Lumipulse p-tau217 assay yields absolute concentrations (pg/ml).

Proteins were manually assigned to biological categories (Inflammatory, Synaptic, Vascular, Metabolic, Neurodegeneration-related, or Amyloid & Tau-related) based on functional annotations from the Human Protein Atlas and hallmarks informed by the manufacturer (Alamar Biosciences). For multifunctional proteins, assignment was based on the predominant biological role most relevant to the central nervous system and neurodegenerative disease.

### Statistical Analyses

Of the 1260 individuals randomized, 1,190 participants were included in the modified intention-to-treat (mITT) population, as previously defined by Ngandu et al^9^. For the present analyses, the study population was restricted to participants included in the mITT population with available baseline NULISA proteomic data (n=1158) to ensure consistency across analyses and facilitate comparability of the results and conclusions (Supplementary Figure 7).

All NULISA proteins plus Lumipulse ptau217 were used for the analyses. For each biomarker and timepoint, values were winsorized at the 1st and 99th percentiles to limit the influence of outliers. NPQ values are already log2-transformed and were therefore not transformed further, whereas Lumipulse p-tau217 was log-transformed. Baseline biomarker levels used as predictors were then standardized to a mean of 0 and a standard deviation of 1 (z-scores).

Baseline characteristics were compared between groups using chi-square tests for categorical variables (sex and *APOE* ε*4 c*arrier status). For continuous variables, independent-samples t-tests were used for baseline and 2-year global cognition, whereas the Mann–Whitney U test was used for other continuous characteristics, according to the distribution of the data as assessed using the Shapiro–Wilk test.

Linear mixed-effects models were fitted using maximum likelihood estimation and included participant-specific random intercepts and slopes for time. Unless otherwise specified, all models were adjusted for age, sex (reference: male), years of education, BMI and study site (reference: Helsinki). Analyses were performed using complete observations for the outcome, predictor and model covariates. For longitudinal cognitive analyses, participants were required to have baseline cognition and at least one cognitive assessment at either the 1- or 2-year follow-up. Two-sided *P* < 0.05 was considered nominally significant. To account for multiple comparisons, *P*-values were adjusted using the Benjamini-Hochberg FDR method and reported when FDR q < 0.05.

To assess intervention-associated changes in biomarkers over time, a linear model was used with outcome the 2-year biomarker change (biomarker level at 2 years) – (biomarker level at baseline)), adjusting for baseline biomarker levels and age, sex, BMI and study site (n = 1,090). Associations between baseline biomarker levels and longitudinal cognitive trajectories were evaluated by linear mixed-effect models, adjusting for randomization group (reference: control), group x time and the prespecified cofactors (age, sex, BMI, study site, education) (formula: ztotal_composite ∼ year x biomarker+ cofactors + year x group + (1 + year | ID); n = 1,146). Cross-sectional associations between baseline biomarker levels and baseline cognition were examined with linear regression model used to evaluate the association between baseline cognition and baseline biomarkers, adjusting for the same cofactors. To investigate whether baseline biomarker levels modified the cognitive response to the intervention, we applied 3-way mixed-effect models with the interaction term baseline biomarker x time x randomization group (formula: ztotal_composite ∼ year x biomarker x group + cofactors+ (1 + year | ID); n = 1,146).

To identify baseline proteins associated with cognitive responder status, a separate analysis was conducted among participants in the intervention group with high adherence (>50% overall adherence across intervention domains or >50% overall adherence in 3 of the domains and 0-50% in the last one; n = 363). Responders were defined as participants showing an improvement in the global cognitive composite score between baseline and the 2-year follow-up (change > 0), whereas participants with no improvement (change ≤ 0) were classified as non-responders. Associations between baseline protein levels and responder status were examined using multivariable logistic regression, adjusting for baseline global cognition, age, sex, education, BMI and study site. Results were expressed as odds ratios (ORs) with 95% confidence intervals (CIs).

Heterogeneity according to APOE4 carrier status and sex was examined using four-way interaction models. APOE4-dependent models included a baseline biomarker x time x randomization group x *APOE ε4* status interaction, whereas sex-dependent models included a baseline biomarker x time x randomization group x sex interaction (n = 1,075). Spearman correlations were calculated between the exploratory NULISA proteins and the core blood-based ADRD biomarkers (Aβ42/40, GFAP, NfL, p-tau181, p-tau217, p-tau231).

Analyses were conducted using R version 4.6.0 (stats package version 4.6.0) for linear regressions and multiple testing correction and lme4 version 2.0.1 for mixed models. Longitudinal biomarker plots were produced using python 3.11 and seaborn 0.13. All other plots were generated in R 4.6.0 using ggplot2.

### Machine learning

All baseline plasma biomarkers measured using the NULISA platform, including established blood-based ADRD biomarkers were included as candidate predictors, together with the clinical covariates used in the mixed-effects models. Individuals with missing cofactors, baseline biomarkers and cognitive scores at baseline and two years were excluded from the analyses (Supplementary Figure 6). An Elastic net regression pipeline was implemented to test whether biomarkers identified through mixed-effects models remained predictive within a multivariable machine learning framework that accounts for correlations between predictors. This approach identifies sets of biomarkers that jointly predict cognitive change over two years, and, separately, biomarkers whose association with cognitive change depends on the intervention group. Elastic net combines L1 (Lasso) and L2 (Ridge) regularization**^25^**, allowing correlated predictors to be retained while shrinking predictors with limited contribution. This property is advantageous for proteomic data, in which correlated biomarkers may reflect overlapping biological pathways.

Analyses were performed in Python 3.11 using ElasticNet from scikit-learn 1.7. The outcome was two-year change in the global cognitive composite score. As Elastic Net penalization is scale-dependent, all predictors were standardized (z-scored) using the mean and SD estimated from the training fold.

Model robustness was assessed using nested cross-validation, with an outer 4-fold loop for performance evaluation and an inner 3-fold loop for hyperparameter tuning. A 4-fold outer split was chosen to preserve adequate per-fold sample size given the limited size of the study cohort, particularly for the control-only and intervention-only analyses.

For each outer training fold, hyperparameters were optimized with a randomized search, by minimizing mean squared error (MSE) on the inner validation folds: the regularization strength (alpha) was searched on a logarithmic scale from 10 to 10², and the L1/L2 mixing ratio (l1_ratio) was searched uniformly between 0.1 and 1.0. Candidate hyperparameter combinations yielding fewer than five non-zero coefficients were excluded from consideration, to avoid overly sparse, unstable models and balance predictive performance against biological interpretability. For each outer fold, one final elastic net model was fit on the full outer training set using the selected hyperparameters, yielding four models per cohort. Predictive performance was evaluated on each corresponding outer test fold using MSE, reported as mean ± SD across the four outer folds.

Predictor robustness was evaluated using two complementary perspectives, computed across the four fold-specific final models: (i) coefficient magnitude, reflecting each biomarker’s direct contribution to predicted cognitive change within a given fold’s model; (ii) shrinkage resistance, defined as the regularization strength (alpha) at which each biomarker’s coefficient first reached zero along the Elastic net regularization path, with higher thresholds indicating greater robustness to penalization and lower likelihood of representing noise. Attention was paid to biomarkers nominally significant in mixed-effects models, repeatedly selected across Elastic Net folds with large coefficient magnitudes and persistent under strong regularization.

Analyses were performed separately in three cohorts: the control cohort (535 individuals), to identify biomarkers predictive of cognitive trajectories in the absence of intervention; the intervention cohort (536 individuals), to identify biomarkers associated with cognitive response under intervention; and the full cohort with biomarker x group interaction terms (1071 of individuals), to identify biomarkers whose association with cognitive change differed between intervention and control groups.

## Results

### Participant characteristics

The present study included all participants from the modified Intention-To-Treat (mITT) FINGER RCT population with available banked samples (N=1158 at baseline, N=1105 at 2 years follow up). The trial population has been described previously^9^. The mean age at baseline was 69.0 (SD 4.7) in the intervention group (N=580) and 68.6 (SD 4.7) in the control group (N=578). There were no significant differences between groups considering age, sex, education, *APOE ε4* carrier status, plasma ptau217, body mass index or global cognition (total composite of a neuropsychological test battery, the primary outcome of the trial) at baseline (all P >0.05; Table 1). None of the plasma proteins differed significantly between the intervention and control groups at baseline (all P >0.05, Supplementary Table 1).

**Table 1.** Cohort characteristics.

| **Baseline characteristics** | **N** | **Total<br>(N=1158)** | **Control<br>(N=578)** | **Intervention<br>(N=580)** | **P-value** |
|---|---|---|---|---|---|
| **Age, years** | 1158 | 68.8 (4.7) | 68.6 (4.7) | 69.0 (4.7) | 0.196 |
| **Women** | 1158 | 540 (46.6%) | 275 (47.6%) | 265 (45.7%) | 0.559 |
| **Education, years** | 1156 | 10.0 (3.4) | 10.0 (3.4) | 10.0 (3.4) | 0.958 |
| **APOE4 carrier** | 1087 | 349 (30.01%) | 181 (31.3%) | 168 (29.0%) | 0.400 |
| **Blood p-tau217, pg/mL** | 1154 | 0.10 [0.08-0.15] | 0.10 [0.08-0.15] | 0.10 [0.08-0.15] | 0.900 |
| **BMI** | 1149 | 28.3 (4.7) | 28.1 (4.8) | 28.4 (4.5) | 0.423 |
| **NTB global cognition** | 1157 | 0.00 (0.57) | 0.02 (0.59) | -0.02 (0.55) | 0.145 |

Data are presented as mean (SD), median [interquartile range] or N (%), as appropriate. P-values compare the intervention and control groups and were calculated using independent-samples t-tests or Mann–Whitney U tests for continuous variables, according to their distribution, and chi-square tests for categorical variables. P-tau217 measured with Lumipulse. BMI, body mass index; NTB, neuropsychological test battery.

### Associations between protein levels and age, sex, *APOE ε4* genotype and cognition

This study included 115 proteins for the relationship with cognition and intervention response and utilized the canonical AD-biomarkers as reference biomarkers (Project overview, Fig. 1). Baseline protein levels showed correlations with age and sex, whereas correlations with *APOE ε4* carriage status were more limited (Supplementary Fig. 1). Among the strongest associations that remained significant after FDR correction, FABP3, FOLR1 and VCAM1 were positively correlated with age (r = 0.25–0.28). NPTX2 levels were lower in women than in men (r = −0.42), whereas SAA1 levels were higher in women (r = 0.33).

**Fig. 1.**
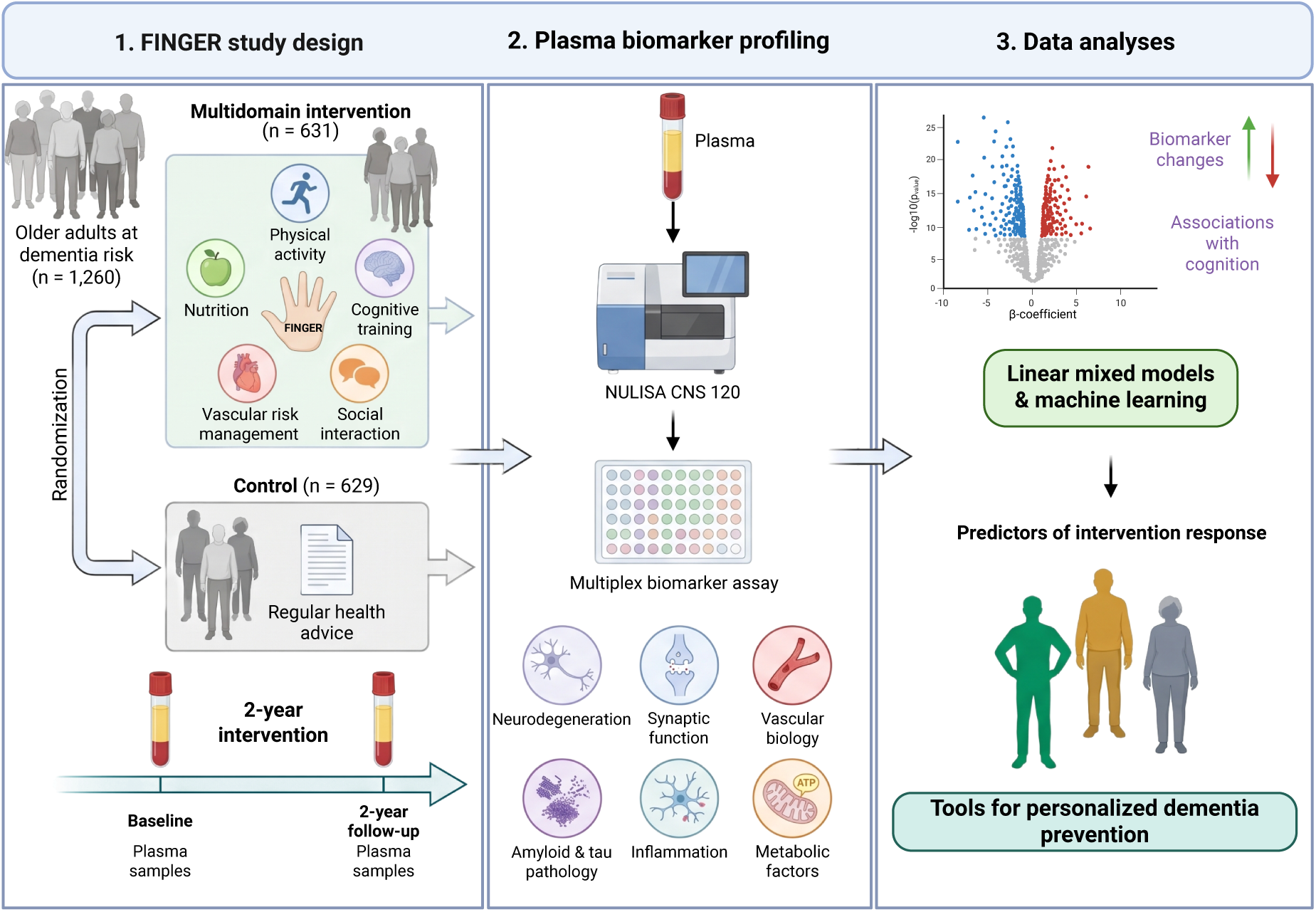
Overview of the FINGER study design, plasma proteomic profiling and analytical framework. The Finnish Geriatric Intervention Study to Prevent Cognitive Impairment and Disability (FINGER) randomized 1,260 older adults at increased risk of dementia to a 2-year multidomain lifestyle intervention (n = 631) or a control group receiving regular health advice (n = 629). The intervention comprised nutritional guidance, physical activity, cognitive training, social interaction and vascular risk management. Plasma samples collected at baseline and the 2-year follow-up were analyzed using the NULISA CNS 120 multiplex proteomic assay, covering proteins involved in neurodegeneration, synaptic function, vascular biology, amyloid and tau pathology, inflammation and metabolism. Statistical modeling and machine-learning approaches were used to investigate intervention-related changes in circulating proteins, associations with cognitive outcomes and baseline protein predictors of cognitive response to the intervention, with the aim of informing personalized dementia prevention strategies. Schematic created in BioRender. Pola, I. (2026). https://BioRender.com/3rgxqsp.

Six biomarkers were nominally associated with cognitive performance at baseline (spanning neurodegeneration-related, inflammatory, vascular and synaptic pathways (P < 0.05; Fig. 2A-B, Supplementary Table 2). Specifically, higher MME levels were associated with better cognitive performance at baseline, whereas higher levels of VGF, UCHL1, VEGFA, NEFH and CHIT1, were associated with poorer baseline cognition.

**Fig. 2.**
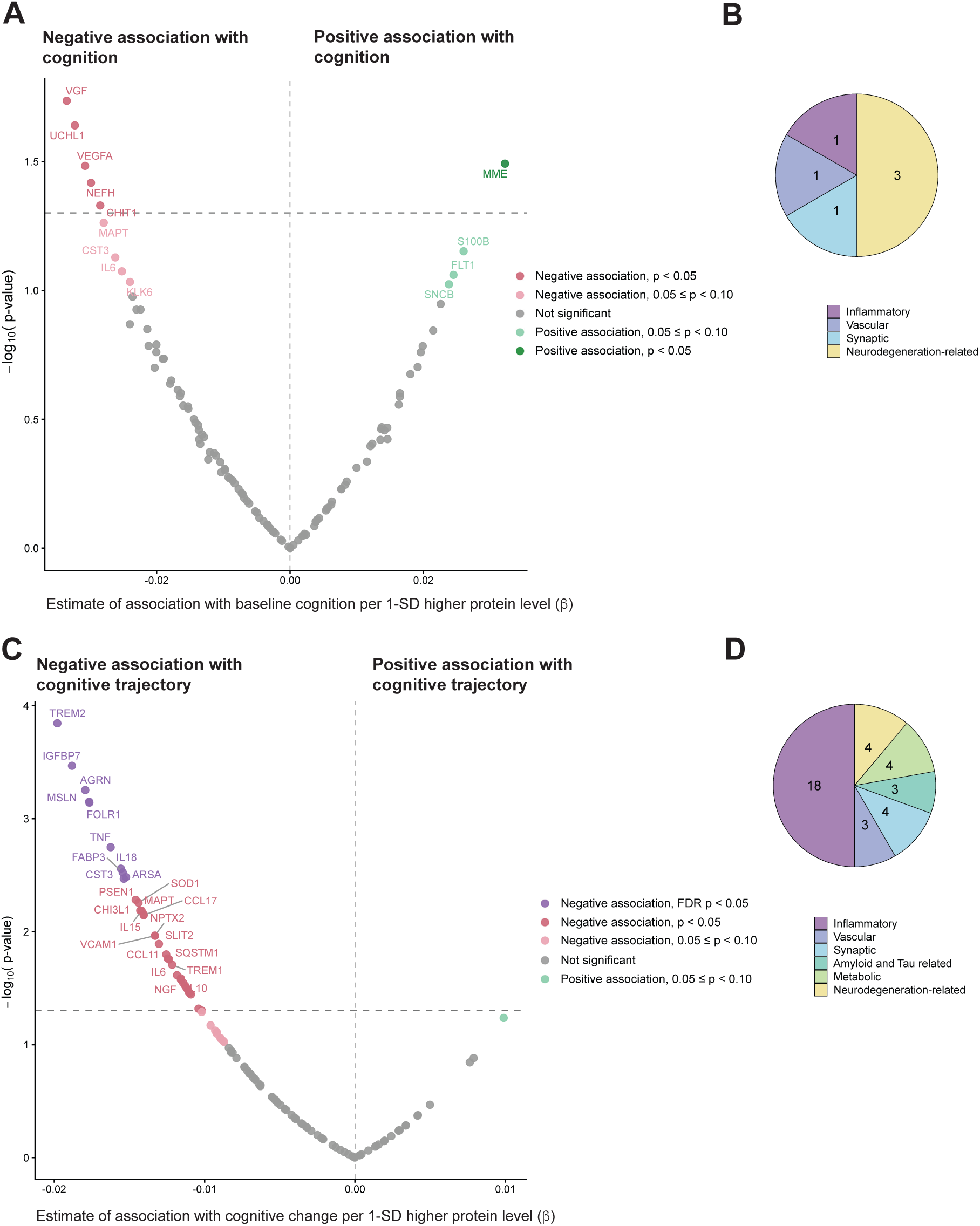
Associations of baseline plasma proteins with cognitive performance and longitudinal change. **A**. Volcano plot showing associations between baseline plasma protein levels and baseline cognitive performance, measured using the Neuropsychological Test Battery (NTB) global composite score, assessed by multivariable linear regression (n = 1,146). **B.** Distribution of proteins nominally associated with baseline cognitive performance (P < 0.05) across biological categories. **C.** Volcano plot showing associations between baseline plasma protein levels and longitudinal changes in the NTB global composite score over the 2-year follow-up, assessed using linear mixed-effects models (n = 1,146). Each point in the volcano plots represents a protein. The x-axis shows the regression coefficient (β) per 1-standard-deviation (SD) increase in baseline protein level, and the y-axis shows −log₁₀(P). Negative β coefficients indicate poorer baseline cognitive performance or less favorable longitudinal cognitive trajectories, whereas positive coefficients indicate better performance or more favorable trajectories. Horizontal dashed lines indicate the nominal significance threshold (P = 0.05). Colors indicate the direction and statistical significance of associations, with FDR-significant associations (FDR P < 0.05) additionally highlighted in panel C. All models were adjusted for age, sex, years of education, body mass index and study site; longitudinal models were additionally adjusted for randomization group its interaction with time. **D.** Distribution of proteins nominally associated with longitudinal cognitive trajectories (P < 0.05) across biological categories.

A total of 35 baseline plasma proteins predicted change in global cognition over time, independently of randomization group, with half of them involved in inflammatory mechanisms (Fig. 2C-D). All significant associations were negative, indicating that higher baseline protein levels were associated with less favorable cognitive trajectories. Of these 35 proteins, 10 remained significant after FDR correction: TREM2, AGRN, FOLR1, IGFBP7, MSLN, TNF, ARSA, CST3, FABP3 and IL18 (FDR-adjusted *P*<0.05; Figure 2C, Supplementary Table 3).

### FINGER intervention effects on protein changes

To determine the effect of the multidomain FINGER intervention on circulating proteins, longitudinal changes were compared between the intervention and control groups. The intervention was associated with changes in plasma levels of eight proteins, at the nominal significance level (*P* < 0.05; Fig. 3A Supplementary Table 4), representing inflammatory (n = 2), vascular (n = 2), amyloid-and tau-related (n = 2), and metabolic (n = 1) categories (Fig. 3B). Compared with the control, participants in the intervention group had greater increases in CRH and FLT1 and a greater decrease in MME (Fig. 3C). IL7, PSEN1 and TREM1 levels increased in both groups, but to a lesser extent in the intervention. In contrast, CD40LG and FGF2 remained relatively stable in the intervention, whereas they were increased in the control group.

**Fig. 3.**
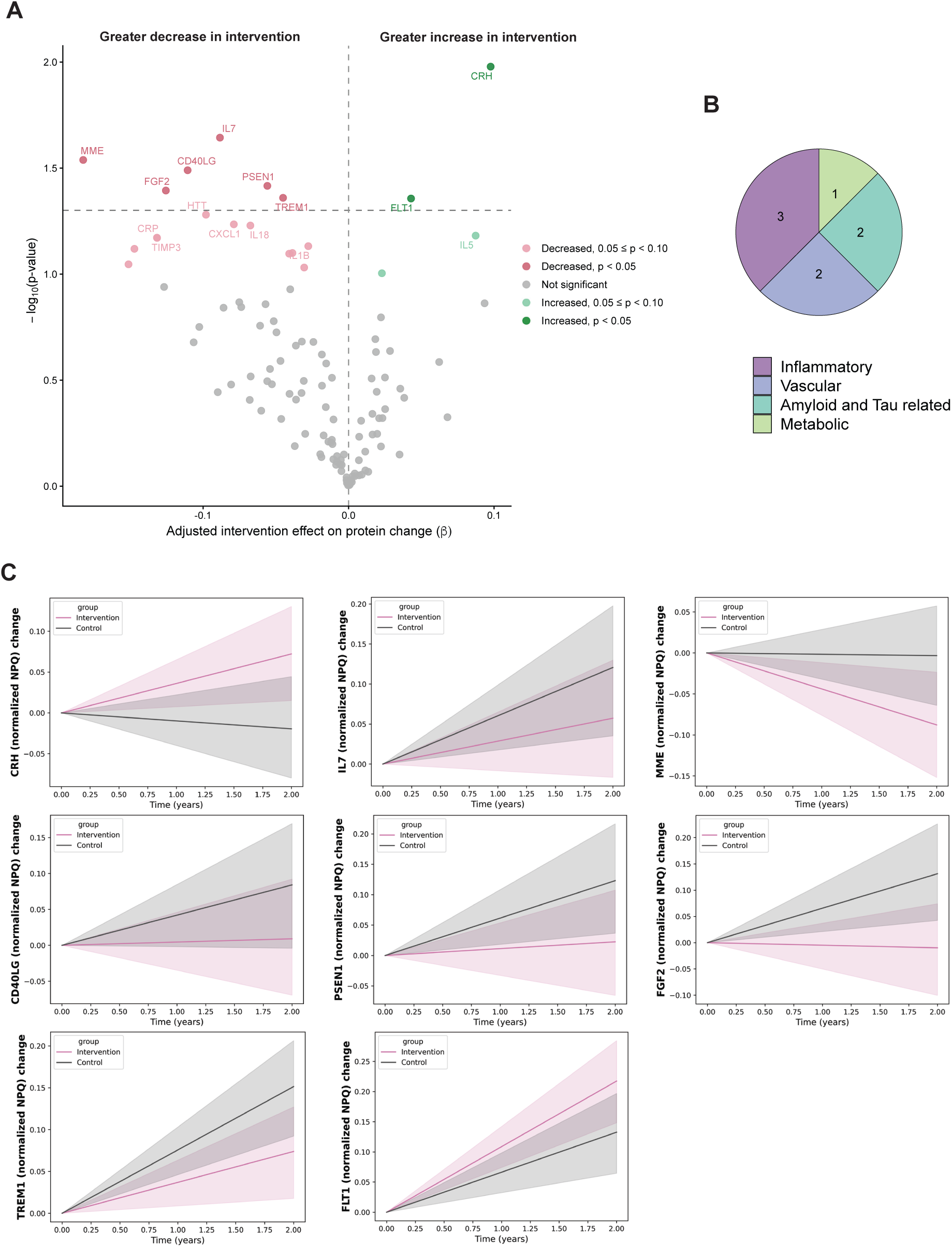
Effects of the FINGER multidomain intervention on circulating plasma protein levels over 2 years. **A.** Volcano plot showing differences in 2-year changes in plasma protein levels between the intervention and control groups (n = 1,093), assessed using multivariable linear regression adjusted for baseline protein levels, age, sex, body mass index and study site. Each point represents a protein. The x-axis shows the adjusted intervention effect (β) on protein change, with positive coefficients indicating greater increases and negative coefficients indicating greater decreases in the intervention group relative to the control group. The y-axis shows −log₁₀(P), and the horizontal dashed line indicates the nominal significance threshold (P = 0.05). Dark green and dark red points indicate proteins showing nominally significant positive and negative intervention effects, respectively (P < 0.05). Light green and light red points indicate suggestive associations (0.05 ≤ P < 0.10), whereas grey points represent non-significant associations. Selected proteins are labelled. **B.** Distribution of the eight proteins showing nominally significant intervention-related changes (P < 0.05) across biological categories. **C.** Changes in normalized protein expression (NPQ) from baseline to the 2-year follow-up for the eight proteins identified in panel A (CRH, IL7, MME, CD40LG, PSEN1, FGF2, TREM1 and FLT1), shown separately for the intervention (pink) and control (grey) groups. Lines represent estimated changes over time, with shaded areas indicating 95% confidence interval.

### Baseline plasma proteins associated with cognitive response to the intervention

To identify candidate biomarkers predictive of intervention responsiveness, we used linear mixed-effects models examining interactions between baseline protein levels, time and randomization group. These models tested whether the association between baseline protein levels and cognitive change over two years differed between the intervention and control groups.

Eleven proteins showed nominally significant three-way interactions (P < 0.05; Fig. 4A; Supplementary Table 5). These proteins were predominantly synaptic (n = 6) and inflammatory (n = 3), with vascular (n = 1) and metabolic (n = 1) proteins also represented (Fig. 4B). Higher baseline levels of eight proteins (KDR, CRH, CCL2, PTN, BDNF, IL7, SNAP25 and NPTX1) were associated with a more favorable cognitive response to the intervention, whereas higher levels of TREM2, SFRP1 and GDNF were associated with a less favorable cognitive response (Fig. 4C). This was particularly pronounced for KDR, for which higher baseline levels were associated with less favorable cognitive trajectories in the control group but were shifted toward more favorable cognitive outcome in the intervention group (Figure 4D). In contrast, higher baseline TREM2 was associated with less cognitive improvement in the intervention relative to controls (Fig. 4D).

**Fig. 4.**
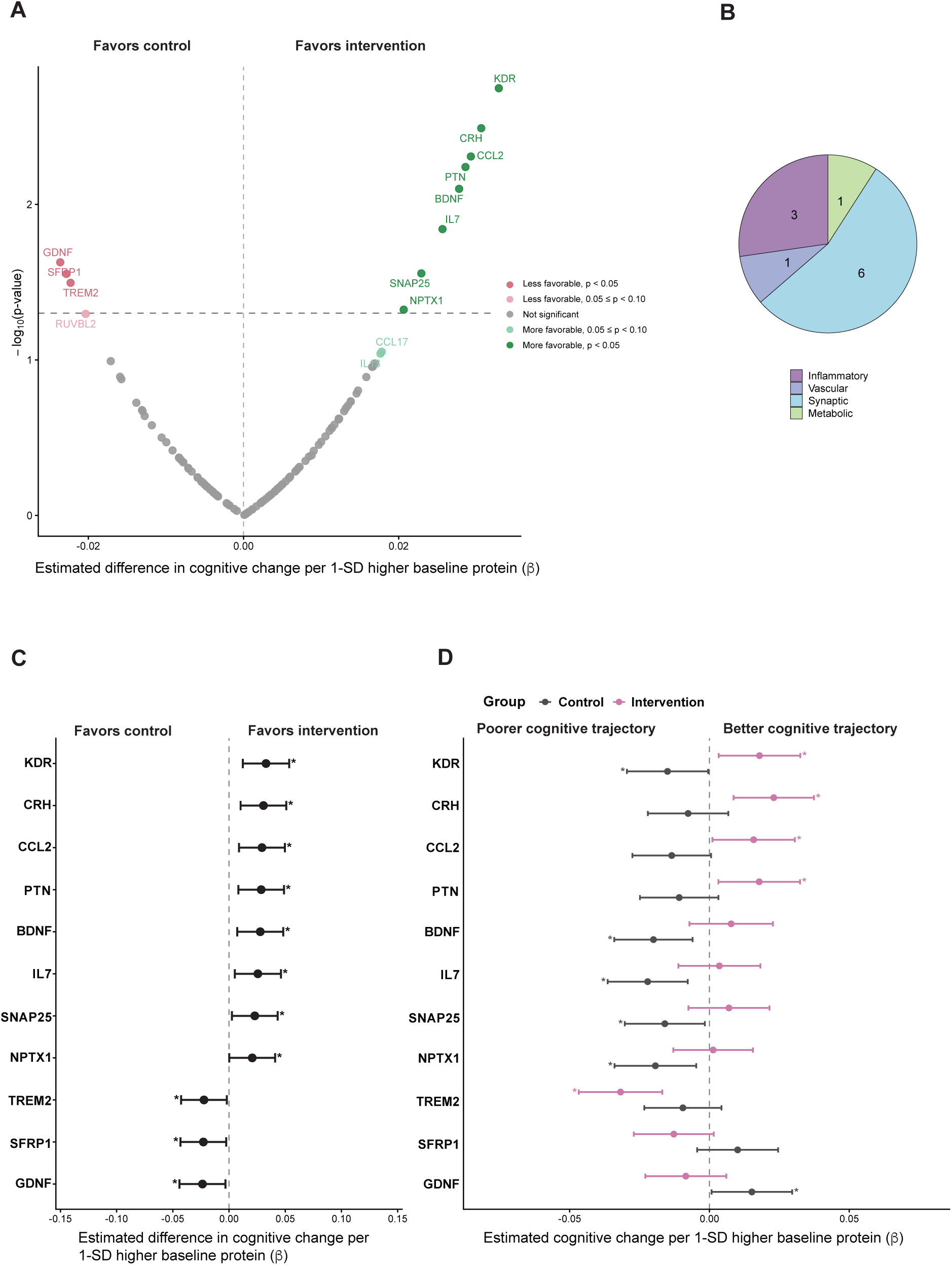
Modification of the FINGER intervention effect on cognitive trajectories by baseline plasma protein levels. **A.** Volcano plot showing the modifying effects of baseline plasma protein levels on intervention-related changes in global cognition, measured using the Neuropsychological Test Battery (NTB) global composite score over the 2-year follow-up (n = 1,146). Associations were assessed using linear mixed-effects models with a baseline protein × time × randomization group interaction. Each point represents a protein. The x-axis shows the estimated difference in cognitive change between the intervention and control groups per 1-standard-deviation (SD) higher baseline protein level (β), and the y-axis shows −log₁₀(P). Positive coefficients indicate a more favorable cognitive response to the intervention relative to control, whereas negative coefficients indicate a less favorable response. Dark green and dark red points indicate nominally significant positive and negative interaction effects, respectively (P < 0.05). Light green and light red points indicate suggestive associations (0.05 ≤ P < 0.10), whereas grey points indicate non-significant associations. The horizontal dashed line denotes the nominal significance threshold (P = 0.05). **B.** Distribution of the eleven proteins showing nominally significant interaction effects (P < 0.05) across biological categories. **C.** Forest plot showing the estimated three-way interaction coefficients (β) and 95% confidence intervals (CIs) for the eleven proteins exhibiting nominally significant modification of the intervention effect on cognitive trajectories. Positive coefficients favor the intervention, whereas negative coefficients favor the control group. **D.** Forest plot showing the associations between baseline protein levels and longitudinal cognitive change separately in the intervention (pink) and control (grey) groups for the eleven proteins identified in panel A. Points represent estimated regression coefficients (β) per 1-SD higher baseline protein level, and horizontal lines indicate 95% CIs. Negative coefficients indicate less favorable cognitive trajectories, whereas positive coefficients indicate more favorable trajectories. All models were adjusted for age, sex, years of education, body mass index and study site. Asterisks in panels C and D indicate nominal statistical significance (P < 0.05).

### *APOE ε4-* and sex-modification of biomarker associations with intervention-related cognitive change

We further investigated whether *APOE ε4* status modified the association between baseline plasma protein levels and the intervention-related cognitive change over two years. Using linear mixed-effects models, we examined four-way interactions between baseline protein levels, time, randomization group and *APOE* ε*4* status. Six proteins showed nominally significant interactions (P < 0.05; Fig. 5A; Supplementary Table 6). These proteins were classified as neurodegeneration-related (n = 3), inflammatory (n = 1), vascular (n = 1), and synaptic (n = 1) (Fig. 5B). For four proteins (HTT, TARDBP, SNCA, NRGN), higher baseline levels were associated with larger intervention effect on cognitive trajectories among *APOE ε4* carriers than non-carriers. In contrast, higher baseline CXCL1 and PDGFRB levels were associated with larger intervention effect among non-carriers than carriers.

**Fig. 5.**
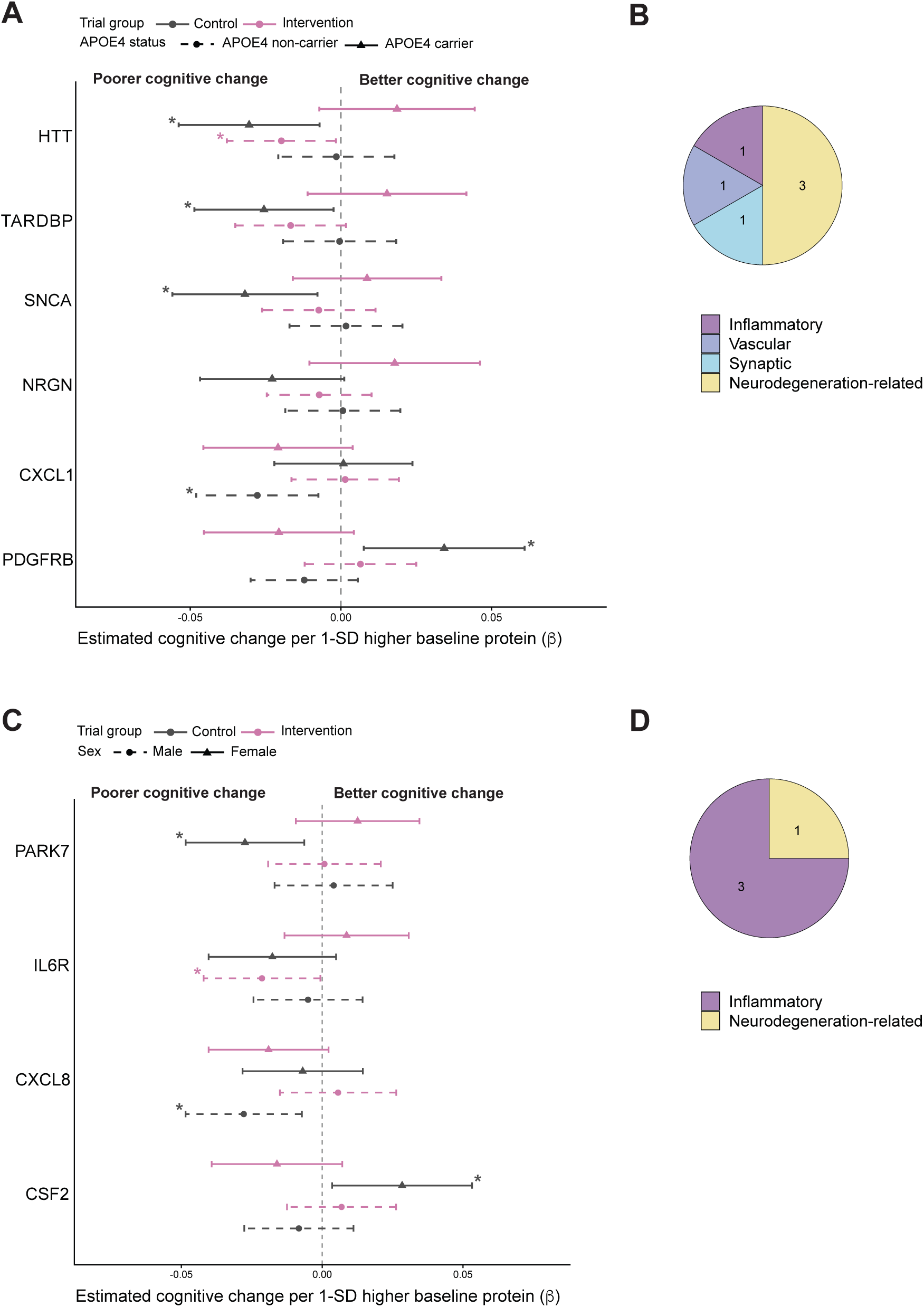
APOE ε4- and sex-dependent modification of FINGER intervention effects by baseline protein levels. **A**. Forest plot showing associations between baseline plasma protein levels and longitudinal changes in global cognition, measured using the Neuropsychological Test Battery (NTB) global composite score, stratified by randomization group and *APOE ε4* carrier status. Six proteins showing nominally significant four-way interactions (baseline protein × time × randomization group × *APOE ε4* status) are presented. **B.** Distribution of these six proteins across biological categories. **C**. Forest plot showing associations between baseline plasma protein levels and longitudinal cognitive change, stratified by randomization group and sex, for four proteins showing nominally significant four-way interactions (baseline protein × time × randomization group × sex). **D.** Distribution of these four proteins across biological categories. In panels A and C, points represent subgroup-specific regression coefficients (β) for cognitive change per 1-standard-deviation (SD) higher baseline protein level, and horizontal lines indicate 95% confidence intervals (CIs). Positive coefficients indicate more favorable cognitive trajectories, whereas negative coefficients indicate less favorable trajectories. Pink and grey indicate the intervention and control groups, respectively. In panel A, solid lines and triangles represent *APOE ε4* carriers, whereas dashed lines and circles represent non-carriers. In panel C, solid lines and triangles represent women, whereas dashed lines and circles represent men. Proteins were selected based on nominally significant four-way interaction effects (P < 0.05). Asterisks indicate nominally significant associations within individual subgroups (P < 0.05). Models were adjusted for age, sex, years of education, body mass index and study site, with sex omitted as a separate covariate in the sex-interaction models. All analyses included 1,075 participants.

Sex-dependent differences were also observed, with four proteins showing nominally significant four-way interactions (P < 0.05; Fig. 5C; Supplementary Table 7). Three proteins were inflammatory (IL6R, CXCL8 and CSF2), and one neurodegeneration-related (PARK7) (Fig. 5D). Higher baseline PARK7 and IL6R levels were associated with more favorable cognitive response to the intervention among women than men. In contrast, higher baseline CXCL8 levels were associated with a more favorable cognitive response to the intervention among men than women, whereas higher CSF2 levels were associated with a less favorable cognitive response to the intervention among women.

### Machine Learning validation of biomarkers predicting intervention-associated cognitive response

In addition to the mixed models, we applied Elastic Net regression with protein x intervention interaction terms as a final analytical step to identify candidate biomarkers predictive of intervention-related cognitive response. The machine-learning models included the established blood-based ADRD biomarkers alongside the broader NULISA biomarker panel (Supplementary Table 1), as candidate predictors, together with clinical and demographic variables. Concordance between biomarkers identified by the two complementary approaches (Elastic Net and mixed-effect models) was used to assess the robustness of our findings. Model development and evaluation were performed using four-fold nested cross-validation, with detailed results presented in Supplementary Results, Supplementary Tables 8–12 and Supplementary Figures 2-4.

In summary, the machine-learning models consistently identified CRH as a potential predictor of intervention response with the CRH × intervention interaction retained in all four cross-validation folds. while PTN × intervention and CCL2 × intervention was also frequently selected, appearing in three of four folds. CRH and CCL2 showed some of the strongest positive interaction effects, suggesting that higher baseline concentrations were associated with greater cognitive benefit from the intervention relative to the control group. Among proteins contributing to change in cognition independently of group assignment, NULISA p-tau217 and SOD1 were consistently selected across all four folds.

Complementary analyses using separate intervention and control models further highlighted CRH, CCL2, and PTN as predictors of cognitive change in the intervention group, with more limited contributions in controls (Supplementary Results). These findings were consistent with the mixed-effects analyses, which identified positive associations between higher baseline levels of CRH, CCL2, and PTN and intervention-related cognitive improvement over time (Suppl. Fig. 5). Additional concordance between the two approaches was observed for KDR, BDNF, GDNF, RUVBL2, and TREM2 (Supplementary Results, Suppl. Fig. 5).

### Baseline plasma proteins associated with cognitive response among highly adherent intervention participants

Complementing the approach of investigating proteins as predictors of response, we compared baseline protein profiles between responders and non-responders among intervention-adherent participants (N=363) aiming to identify biological mechanisms associated with cognitive benefit from the multidomain lifestyle intervention.

Among the adherent individuals, 283 (78.0%) were classified as responders, defined by a positive two-year change in the global cognitive composite, and 80 (22.0%) as non-responders. Responders showed a mean cognitive improvement of 0.384 (SD 0.231), whereas non-responders exhibited a mean decline of −0.218 (SD 0.164) over two years (Fig. 6A; Supplementary Table 13). Responders were younger than non-responders (68.1 versus 69.8 years; P = 0.005), whereas no significant differences were observed in sex, education, BMI, *APOE ε4* status or baseline global cognition (Supplementary Table 13).

**Fig. 6.**
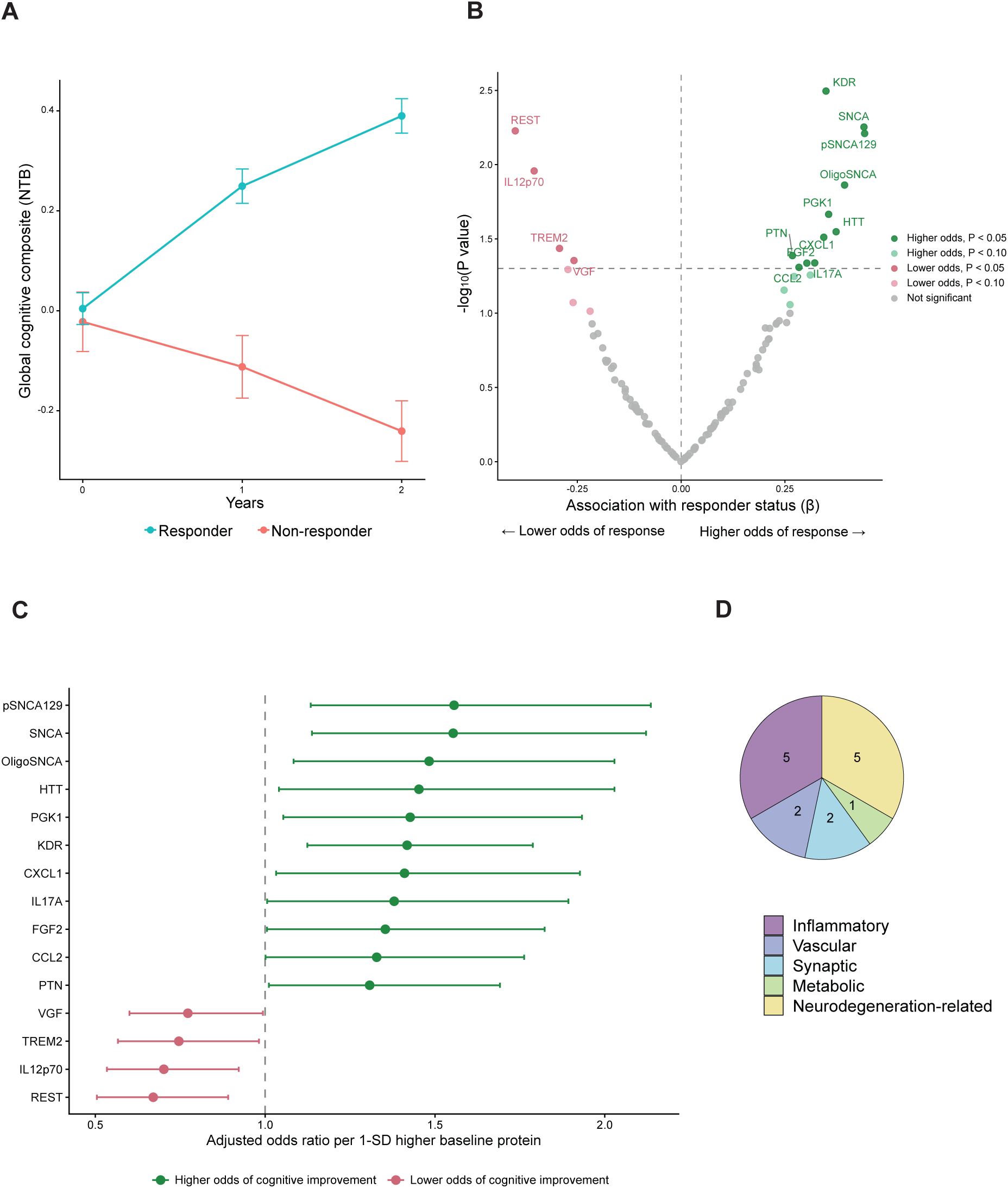
Baseline plasma proteins as predictors of cognitive response among highly adherent FINGER intervention participants. **A.** Observed mean global cognitive performance, measured using the Neuropsychological Test Battery (NTB) global composite score, at baseline, 1 year and 2 years among cognitive responders (n = 283) and non-responders (n = 80) with high adherence to the FINGER intervention. Responders were defined as participants showing an improvement in the NTB global composite score between baseline and 2 years (change > 0), whereas non-responders showed no improvement (change ≤ 0). Points represent observed group means, and error bars indicate the standard error of the mean (SEM). **B.** Volcano plot showing associations between baseline plasma protein levels and cognitive responder status, assessed using multivariable logistic regression. Each point represents a protein. The x-axis shows the regression coefficient (β) for responder status per 1-standard-deviation (SD) higher baseline protein level, and the y-axis shows −log₁₀(P). Positive coefficients indicate higher odds of cognitive response, whereas negative coefficients indicate lower odds. Dark green and dark red points indicate nominally significant associations with higher and lower odds of response, respectively (P < 0.05). Light green and light red points indicate suggestive associations (0.05 ≤ P < 0.10), whereas grey points indicate non-significant associations. The horizontal dashed line indicates the nominal significance threshold (P = 0.05). **C.** Forest plot showing adjusted odds ratios (ORs) and 95% confidence intervals (CIs) for the 15 proteins nominally associated with cognitive responder status (P < 0.05). ORs are expressed per 1-SD higher baseline protein level. Green and red indicate associations with higher and lower odds of cognitive response, respectively. The vertical dashed line indicates no association (OR = 1). **D.** Distribution of the 15 proteins nominally associated with cognitive responder status across biological categories. Logistic regression models were adjusted for baseline NTB global composite score, age, sex, years of education, body mass index and study site. All protein associations shown in panel C were nominally significant (P < 0.05).

Fifteen baseline proteins were nominally associated with cognitive responder status (P < 0.05; Fig. 6B, C; Supplementary Table 14). These proteins represented inflammatory (n = 5), neurodegeneration-related (n = 5), synaptic (n = 2), vascular (n = 2) and metabolic categories (n = 1) (Fig. 6D). Higher baseline levels of 11 proteins (KDR, SNCA, pSNCA129, OligoSNCA, PGK1, HTT, CXCL1, PTN, FGF2, IL17A and CCL2) were associated with greater odds of cognitive response. Among the strongest associations were SNCA (OR = 1.55, P = 0.006), pSNCA129 (OR = 1.56, P = 0.006) and KDR (OR = 1.41, P = 0.003). Conversely, higher baseline levels of REST, IL12p70, TREM2 and VGF were associated with lower odds of cognitive response.

### Correlations between baseline exploratory proteins and established plasma ADRD biomarkers

Widespread correlations were observed between baseline exploratory proteins and established ADRD biomarkers, with numerous correlations remaining significant after FDR correction (Suppl. Fig. 6). Focusing on seven proteins showing convergent associations with intervention-related cognitive trajectories in the three-way interaction and machine-learning analyses, CCL2 and BDNF were negatively correlated with Aβ42/40 (r = −0.27 and −0.29, respectively), whereas PTN showed a positive correlation with p-tau217 (r = 0.16). CRH showed weak positive correlations with Aβ42/40 (r = 0.08) and GFAP (r = 0.07), while TREM2 was positively correlated with NfL (r = 0.21) and GFAP (r = 0.16). In contrast, KDR and GDNF showed no FDR-significant correlations with the reference biomarkers.

Among the fifteen proteins associated with cognitive responder status among highly adherent intervention participants, REST showed positive correlations with p-tau181 (r = 0.37), p-tau231 (r = 0.34) and p-tau217 (r = 0.28), whereas SNCA and pSNCA129 were negatively correlated with Aβ42/40 (r = −0.25 for both). Of the eight proteins showing intervention-related changes, PSEN1 was positively correlated with phosphorylated tau biomarkers (r = 0.18–0.20) and NfL (r = 0.20), whereas CD40LG and IL7 were negatively correlated with Aβ42/40 (r = −0.23 and −0.20, respectively). Finally, among 35 proteins associated with longitudinal cognitive trajectories, MAPT and FABP3 had strong positive correlations with phosphorylated tau biomarkers, while FOLR1 was positively correlated with NfL (r = 0.46) and GFAP (r = 0.41).

## Discussion

The future of dementia prevention will likely require approaches that can be tailored to individuals’ biological profiles and possibly integrated with disease-modifying therapies. FINGER, the first large multidomain lifestyle trial to demonstrate cognitive benefits in older adults at increased risk of dementia, provides a unique opportunity to investigate the molecular correlates of intervention response. Through longitudinal profiling of a panel of exploratory blood proteins quantified before and after the two-year intervention period, we identified changes in proteins involved in inflammation, synaptic integrity, pathological protein aggregation, vascular- and metabolic-processes with greatest effects on CRH, IL7 and MME. Several baseline proteins modified the cognitive response to the FINGER intervention, suggesting biological determinants of intervention responsiveness. Additional proteins exhibited *APOE* ε*4*- or sex-dependent effect modification, revealing heterogeneity in the biological mechanisms associated with cognitive response. Several proteins associated with intervention-related outcomes showed significant correlations with established ADRDs, although these associations were generally modest, suggesting that the biological pathways they reflect may partly overlap with, but are not fully captured by traditional ADRD biomarkers. Supporting these findings, machine learning analyses integrating multiple biomarkers and participant characteristics identified multivariable signatures predictive of intervention benefit. Finally, comparison of baseline biomarker profiles between intervention-adherent responders and non-responders provided further biological insight into pathways that may contribute to favorable cognitive outcomes following multidomain lifestyle intervention.

Given the limited availability of accessible biomarkers for predicting cognitive decline in older adults without dementia, we first examined associations between baseline plasma proteins and longitudinal cognition in the total FINGER cohort, adjusting for randomization group. We found that a set of proteins predicted less favorable longitudinal cognitive outcomes independent of intervention assignment. These markers are related to immune- and inflammatory processes ^26^, neurodegeneration pathways, synaptic integrity ^27,28^, and metabolic regulation ^29^. The broad range of biological processes captured by these proteins suggests that cognitive change is influenced by multiple interconnected mechanisms linked to dementia risk and age-related decline. Several of these proteins, including TREM2, IGFBP7, and IL18, have previously been associated with long-term cognitive decline and dementia risk in proteomic studies, lending further support to their relevance as early markers of cognitive aging and neurodegeneration.^30–33^.

Biomarkers associated with cognitive decline are not necessarily the same as those mediating intervention-related cognitive benefits. In a separate study, we investigated effects of the FINGER intervention on longitudinal changes in established ADRD blood biomarkers, including p-tau, Aβ42/40, GFAP and NfL^17^ and found no effects on these markers. Within the broader CNS disease proteomic panel, the present study however identified nominal intervention-related changes in several proteins, particularly CRH (corticotropin releasing hormone) and MME (membrane metallo-endopeptidase, commonly known as neprilysin). In the brain, MME is one of the most significant amyloid-β (Aβ) degrading enzymes and known to decline with age and across several neurodegenerative disorders^34^. Levels of MME in the periphery, however, are generally elevated in association with poor metabolic health^35^ consistent with its role in the regulation of multiple metabolic and vascular proteins including incretins (e.g. glucagon-like peptide-1, GLP-1^36^). Thus, the intervention-related reduction in circulating MME may reflect favorable changes in peripheral metabolic regulation rather than diminished cerebral Aβ clearance. CRH was found to be increased by the FINGER intervention. CRH is produced in the hypothalamus where it induces release of corticosteroids^37,38^. This stress related hormone has previously been shown to be decreased in plasma across several age-related neurodegenerative disorders^39,40^ and in postmortem brain material from AD patients^41,42^, plausibly resulting from a general endocrine insufficiency and/or a negative feedback loop from cortisol over-activation. Furthermore, CRH is increased with physical exercise and with stress^43^, and is also linked with inflammation, energy homeostasis and metabolism^44^. The observed increase in circulating CRH may therefore reflect an adaptation in the participants’ general metabolic and immune health, in alignment with the FINGER model. However, increased activation of stress-response pathways remains an alternative explanation, and further investigation is needed to establish the physiological significance of these protein changes. Thus, these results suggest that the early biological effects of multidomain lifestyle intervention may involve neuroendocrine, immune and metabolic processes not adequately captured by conventional markers of amyloid and tau pathology, at least over a two-year period in older adults at increased dementia risk.

Several baseline proteins modified the association between the intervention and subsequent cognitive trajectories, suggesting that the cognitive response to multidomain intervention may differ according to the underlying biological profile. Kinase insert domain receptor (KDR/VEGFR2), a key receptor involved in angiogenesis and vascular function^45^, was the strongest effect modifier among the proteins analysed, with higher baseline levels predicting more cognitive improvement under intervention. Interestingly, KDR peripheral levels were previously found lower in individuals with dementia compared to healthy controls, and decreased mRNA levels were observed in the brain of AD mouse models^46^. This may suggest that differences in vascular and angiogenic signalling are relevant to the capacity to respond to lifestyle modification. Several other identified effect modifying proteins were related to synaptic function and neurotrophic signalling (PTN, NPTX1, BDNF, GDNF and SNAP25)^47–49^ or immune and inflammatory processes (CCL2, TREM2 and IL7)^50^. Importantly, the direction of these associations differed across proteins, suggesting that both baseline vulnerability and resilience may shape the potential for intervention benefit. Some protein profiles may reflect biological vulnerability that remains modifiable through lifestyle intervention, whereas others may indicate more established processes with less potential for modification, or conversely a more resilient state with less room for additional benefit. This was apparent among neurotrophic proteins: the unfavourable association of higher BDNF with cognitive trajectories in controls was attenuated under intervention, whereas GDNF showed a more favourable association in controls but a less favourable pattern in intervention participants. In contrast, higher baseline TREM2 was associated with poorer cognitive trajectories in both groups, with a more pronounced negative association under intervention. Given the role of TREM2 in microglial responses and AD-related neurodegeneration^51^, higher circulating levels may identify a state of greater underlying disease or immune-related vulnerability that is less amenable to modification through lifestyle intervention. CRH also emerged as an effect modifier, with higher baseline levels associated with a more favourable cognitive trajectory under intervention. Whereas our previous FINGER analysis found no significant effect modification by established plasma ADRD biomarkers^17^, the present study highlights candidate synaptic, vascular, immune and neuroendocrine pathways that may contribute to heterogeneity in cognitive responses to multidomain lifestyle intervention.

The heterogeneous associations observed across these proteins suggest that intervention responsiveness may depend on multiple interacting biological processes rather than individual biomarkers in isolation. To explore this possibility, we complemented the mixed-effects analyses with machine-learning approaches integrating the broader proteomic panel, established ADRD biomarkers and participant characteristics. P-tau217 was one of the most consistently selected predictors of cognitive change, supporting its established prognostic relevance^52^. In contrast, CRH, CCL2 and PTN showed more intervention-dependent patterns, contributing prominently to predictions in the intervention group but less consistently in controls. Their interaction terms were also repeatedly retained in full-cohort models, providing complementary support for the effect modification observed in the mixed-effects analyses. Together, these findings suggest that CRH, CCL2 and PTN warrant further investigation as candidate biomarkers of differential intervention responsiveness, whereas p-tau217 may primarily capture broader cognitive vulnerability.

Non-modifiable individual characteristics may influence the associations between biomarkers and responsiveness to lifestyle interventions. We therefore, investigated whether baseline plasma proteins modified the FINGER intervention on cognitive trajectories according to *APOE ε4* carrier status and sex. These analyses identified additional effect modifiers particularly in *APOE ε4* carriers. Higher baseline levels of proteins related to proteinopathies (HTT, TARDBP and SNCA) were associated with less favorable cognitive change in the control group, an effect that was neutralized by the intervention. These biomarkers were highly inter-correlated which could potentially be due to an overlapping biology. The *APOE ε4* variant has been recognized as a mediator of "systemic biological vulnerability" across neurodegenerative disorders by altering inflammatory profiles^53^ and impairing neuronal protein degradation systems^54,55^. In this context, our findings indicate that multidomain intervention may partly counteract cognitive vulnerability associated with these processes in *APOE ε4* carriers, offering insights for future tailoring of interventions according to genetic risk. A recent meta-analysis has also shown that *APOE ε4* carriers benefit more from lifestyle-based dementia risk reduction strategies^56^ which may at least partially be due to the herein identified effects. Given their higher risk of dementia and potential side effects from anti-amyloid treatment, *APOE ε4* carriers represent a prioritized group which could be particularly relevant for future precision prevention strategies.

The impact of sex on effect modification revealed four biomarkers with differential associations with cognitive response between men and women. In women, the intervention attenuated a negative association of PARK7 (DJ-1) and a positive association of CSF2 with cognitive change. In men, CXCL8 was associated with poorer cognitive trajectories in the control group, whereas IL6R was positively associated with cognitive change. Three out of four proteins were inflammatory; however no sex differences were observed for the many other inflammatory proteins assessed, suggesting that there is not a general sex-dependent inflammatory-related effect modification. In future analyses, a broader range of biomarkers may provide additional insight into potential sex-dependent differences.

To further characterize the biological profiles associated with favourable cognitive outcomes, we compared baseline plasma proteins between cognitive responders and non-responders among highly adherent FINGER intervention participants. Higher baseline levels of KDR, BDNF and PTN were associated with greater odds of cognitive improvement, potentially implicating vascular and neurotrophic pathways in cognitive outcomes following multidomain lifestyle intervention. Immune-related proteins showed contrasting associations, with higher TREM2 and IL12p70 associated with lower odds of improvement, whereas CCL2, CXCL1 and IL17A showed positive associations. This suggests that distinct immune pathways, rather than a uniform inflammatory profile, may be relevant to cognitive response. Higher baseline REST was also associated with lower odds of improvement. REST has been implicated in neuronal stress resistance and neuroprotection^57^ and shown to be increased following mindfulness-based stress reduction^58^. In our cohort, however, higher REST correlated with older age and several established ADRD biomarkers, despite showing no significant associations with cognitive trajectories. These findings raise the possibility that circulating REST reflects age-related biological vulnerability or compensatory stress responses, although its physiological significance remains uncertain.

This study has several strengths. It represents a unique investigation of circulating proteomic markers within a randomized controlled multidomain lifestyle intervention. The relatively large and deeply phenotyped FINGER cohort, together with longitudinal cognitive assessments, allowed us to examine both changes in biomarkers following the intervention and their associations with cognitive trajectories. The use of highly sensitive NULISA technology enabled the measurement of a broad range of proteins, including low-abundance circulating proteins that may not be captured by conventional proteomic approaches. Finally, the combination of mixed-effects models and Elastic Net provided two complementary approaches to identify intervention-related biomarker signals, while nested cross-validation was used to improve the robustness of the machine-learning analyses.

Some limitations should also be considered. First, although the FINGER trial included a relatively large cohort, statistical power to detect protein–intervention interactions was limited, particularly for the more complex four-way interactions involving *APOE* ε4 status or sex. These exploratory analyses were not based on prespecified protein-specific hypotheses, and their interpretation is challenging given the number of comparisons and potential instability of higher-order interaction estimates. Several of the findings, particularly from the intervention and interaction analyses, did not survive correction for multiple testing and should therefore be considered exploratory and require replication. Although Elastic Net provided a complementary way to assess the consistency of the biomarker signals identified in the mixed-effects analyses, evidence of overfitting and the absence of an external validation cohort, limit the generalizability of the findings. Second, the FINGER eligibility criteria selected older adults at increased dementia risk with cognitive performance within a relatively restricted range, potentially limiting the generalizability of the findings and influencing cross-sectional associations between proteins and cognition. Furthermore, the responder analyses were restricted to highly adherent intervention participants. As adherence may be associated with participant characteristics influencing cognitive outcomes, these results should be interpreted cautiously. Finally, the relatively homogeneous Finnish study population may limit generalizability to more ethnically and socioeconomically diverse populations. Future studies should prioritize adequately powered, hypothesis-driven analyses and independent validation of candidate biomarkers and predictive models across diverse populations.

## Conclusions

Accessible biomarkers that can predict cognitive responses to lifestyle interventions and guide future combination trials are needed to advance precision dementia prevention. The current findings highlight neuroendocrine, vascular, neurotrophic and immune pathways associated with intervention effects and cognition, extending beyond those captured by established ADRD biomarkers. Although independent validation is needed, these results identify molecular candidates that may inform participant selection, treatment stratification, and future trials combining lifestyle and pharmacological interventions.

## Supporting information

Supplementary material - Elastic net

## Data availability statement

FINGER dataset can be requested from University of Eastern Finland. De-identified data can be shared with academic researchers who provide a research plan accepted by the study group (estimated time frame one month). Data transfer agreements must be completed before accessing the data. Enquiries should be directed to.

