## Supplementary figures for "Plasma proteomics reveals signatures of cognitive aging and response to the FINGER multidomain lifestyle intervention"

**Supplementary Figure 1. Correlations of baseline plasma biomarkers with age, sex and APOE ε4 carrier status.** Heatmap showing Spearman correlations between baseline plasma biomarker levels and age, sex and APOE ε4 carrier status. Positive correlations with sex indicate higher biomarker levels in females and negative correlations indicate higher levels in males, with male sex used as the reference. Positive correlations with APOE ε4 status indicate higher biomarker levels in APOE ε4 carriers than in non-carriers. Color intensity represents the Spearman correlation coefficient (ρ), ranging from −1 to 1. Asterisks indicate statistical significance: **FDR P* < 0.05, ***FDR P* < 0.01 and *FDR* ****P* < 0.001.

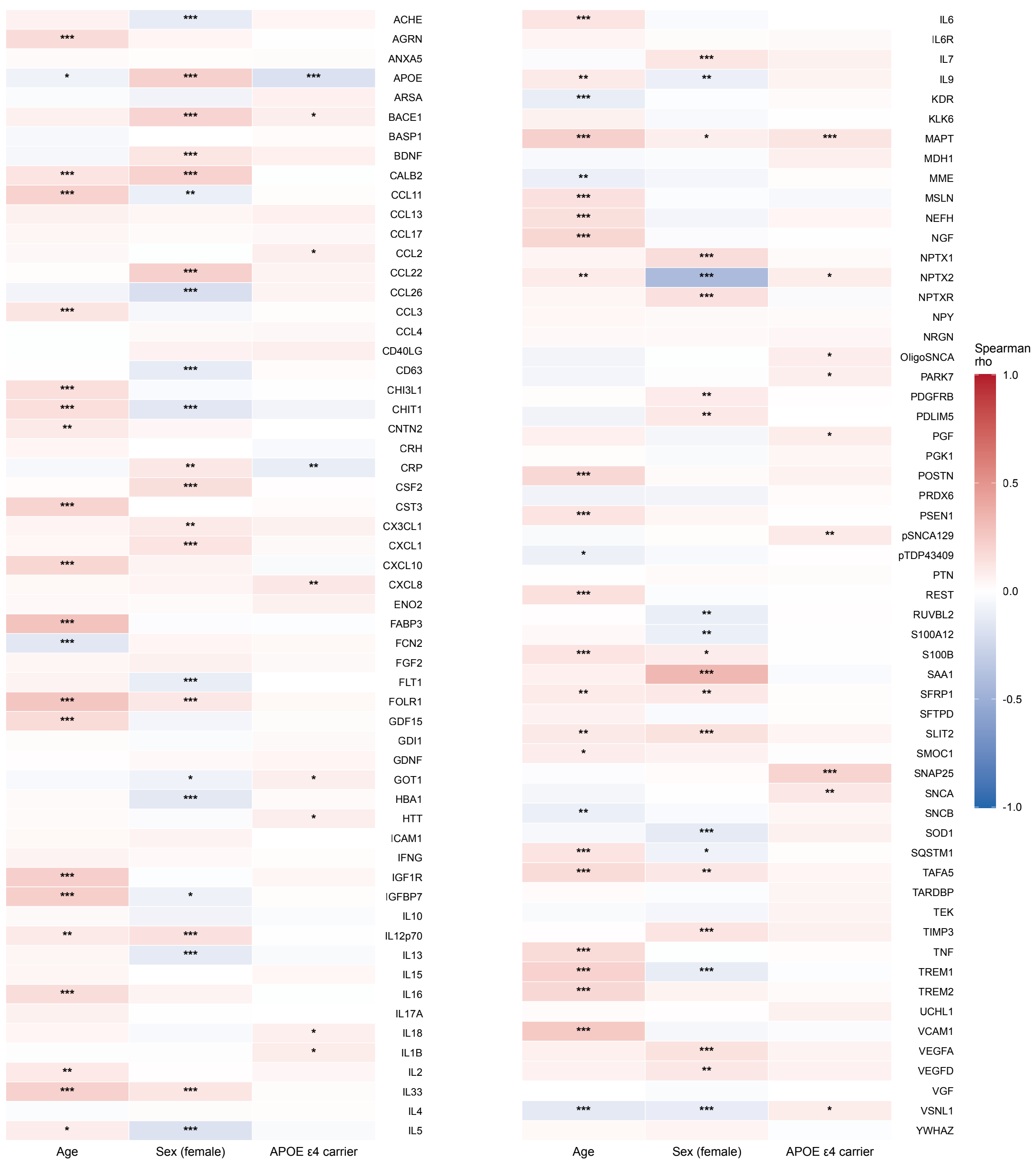

**Supplementary Figure 2. Elastic Net feature selection and coefficient stability in the full cohort with biomarker × intervention-group interaction terms. A-D.** Elastic Net coefficient paths for the four outer cross-validation folds. Each line shows the coefficient of one biomarker, biomarker × group interaction term, or covariate across increasing regularization strength (alpha). Only features retained in the final model for the corresponding fold are shown. For each fold, the best model is represented by the dashed green line. **E.** Coefficients in the best models (one per outer fold) for each feature appearing in at least two of the best models for the full cohort with interaction terms. Red crosses indicate the coefficient estimated in each fold, and *n* indicates selection frequency across the four models (*n* = 4 denotes selection in all four models). Numbers associated with individual coefficients indicate the within-fold rank of each feature according to resistance to penalization, with higher resistance indicating persistence of the feature coefficient under stronger regularization.

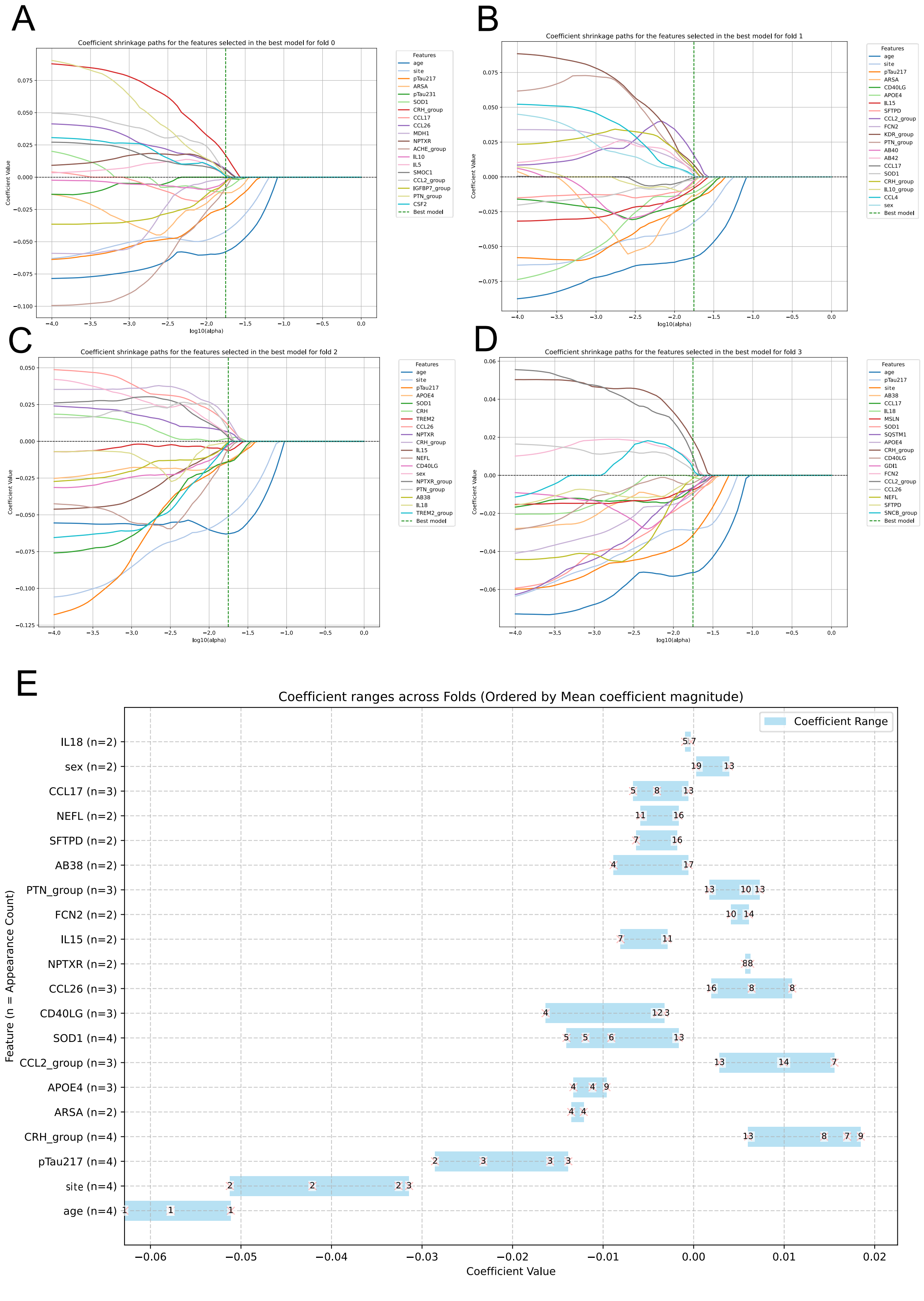

A
Coefficient shrinkage paths for the features selected in the best model for fold 0
Coefficient Value
log10(alpha)
Features
age
site
pTau217
ARSA
pTau231
SOD1
CRH_group
CCL17
CCL26
MDH1
NPTXR
ACHE_group
IL10
IL5
SMOC1
CCL2_group
IGFBP7_group
PTN_group
CSF2
Best model
B
Coefficient shrinkage paths for the features selected in the best model for fold 1
Coefficient Value
log10(alpha)
Features
age
site
pTau217
ARSA
CD40LG
APOE4
IL15
SFTPD
CCL2_group
FCN2
KDR_group
PTN_group
AB40
AB42
CCL17
SOD1
CRH_group
IL10_group
CCL4
sex
Best model
C
Coefficient shrinkage paths for the features selected in the best model for fold 2
Coefficient Value
log10(alpha)
Features
age
site
pTau217
APOE4
SOD1
CRH
TREM2
CCL26
NPTXR
CRH_group
IL15
NEFL
CD40LG
sex
NPTXR_group
PTN_group
AB38
IL18
TREM2_group
Best model
D
Coefficient shrinkage paths for the features selected in the best model for fold 3
Coefficient Value
log10(alpha)
Features
age
pTau217
site
AB38
CCL17
IL18
MSLN
SOD1
SQSTM1
APOE4
CRH_group
CD40LG
GDI1
FCN2
CCL2_group
CCL26
NEFL
SFTPD
SNCB_group
Best model
E
Coefficient ranges across Folds (Ordered by Mean coefficient magnitude)
Coefficient Range
Feature (n = Appearance Count)
IL18 (n=2)
sex (n=2)
CCL17 (n=3)
NEFL (n=2)
SFTPD (n=2)
AB38 (n=2)
PTN_group (n=3)
FCN2 (n=2)
IL15 (n=2)
NPTXR (n=2)
CCL26 (n=3)
CD40LG (n=3)
SOD1 (n=4)
CCL2_group (n=3)
APOE4 (n=3)
ARSA (n=2)
CRH_group (n=4)
pTau217 (n=4)
site (n=4)
age (n=4)
Coefficient Value

**Supplementary Figure 3. Elastic Net feature selection and coefficient stability in the control cohort. A-D**. Elastic Net coefficient paths for the four outer cross-validation folds. Each line shows the coefficient of one biomarker, or covariate across increasing regularization strength (alpha). Only features retained in the final model for the corresponding fold are shown. For each fold, the best model is represented by the dashed green line. **E**. Coefficients in the best models (one per outer fold) for each feature appearing in at least two of the best models for the control cohort. Red crosses indicate the coefficient estimated in each fold, and *n* indicates selection frequency across the four models ($n = 4$ denotes selection in all four models). Numbers associated with individual coefficients indicate the within-fold rank of each feature according to resistance to penalization, with higher resistance indicating persistence of the feature coefficient under stronger regularization.

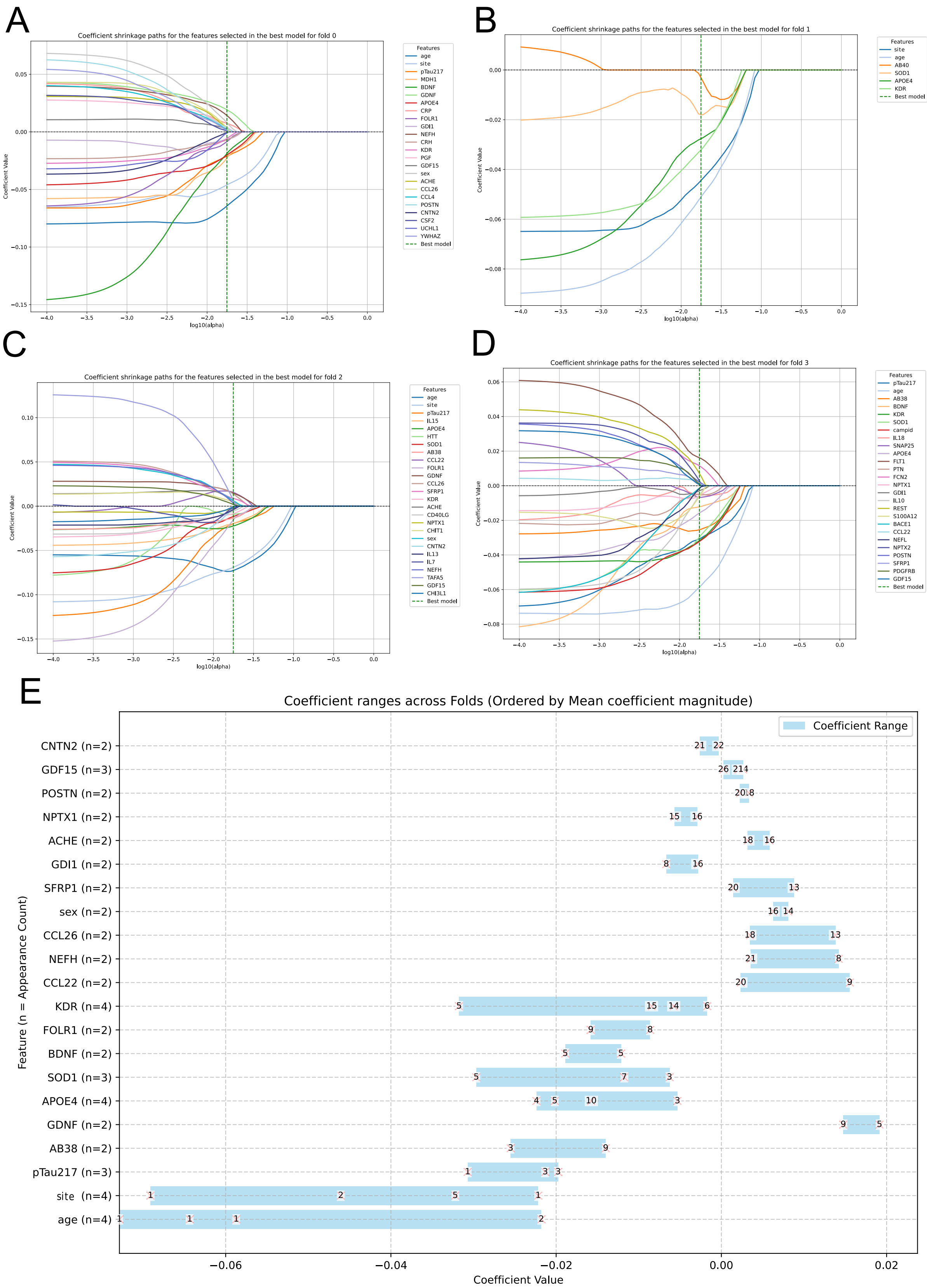
A
Coefficient shrinkage paths for the features selected in the best model for fold 0
Coefficient Value
log10(alpha)
Features
age
site
pTau217
MDH1
BDNF
GDNF
APOE4
CRP
FOLR1
GDI1
NEFH
CRH
KDR
PGF
GDF15
sex
ACHE
CCL26
CCL4
POSTN
CNTN2
CSF2
UCHL1
YWHAZ
Best model
B
Coefficient shrinkage paths for the features selected in the best model for fold 1
Coefficient Value
log10(alpha)
Features
site
age
AB40
SOD1
APOE4
KDR
Best model
C
Coefficient shrinkage paths for the features selected in the best model for fold 2
Coefficient Value
log10(alpha)
Features
age
site
pTau217
IL15
APOE4
HTT
SOD1
AB38
CCL22
FOLR1
GDNF
CCL26
SFRP1
KDR
ACHE
CD40LG
NPTX1
CHIT1
sex
CNTN2
IL13
IL7
NEFH
TAFA5
GDF15
CHI3L1
Best model
D
Coefficient shrinkage paths for the features selected in the best model for fold 3
Coefficient Value
log10(alpha)
Features
pTau217
age
AB38
BDNF
KDR
SOD1
campid
IL18
SNAP25
APOE4
FLT1
PTN
FCN2
NPTX1
GDI1
IL10
REST
S100A12
BACE1
CCL22
NEFL
NPTX2
POSTN
SFRP1
PDGFRB
GDF15
Best model
E
Coefficient ranges across Folds (Ordered by Mean coefficient magnitude)
Coefficient Range
CNTN2 (n=2)
GDF15 (n=3)
POSTN (n=2)
NPTX1 (n=2)
ACHE (n=2)
GDI1 (n=2)
SFRP1 (n=2)
sex (n=2)
CCL26 (n=2)
NEFH (n=2)
CCL22 (n=2)
KDR (n=4)
FOLR1 (n=2)
BDNF (n=2)
SOD1 (n=3)
APOE4 (n=4)
GDNF (n=2)
AB38 (n=2)
pTau217 (n=3)
site (n=4)
age (n=4)
Feature (n = Appearance Count)
Coefficient Value

**Supplementary Figure 4. Elastic Net feature selection and coefficient stability in the intervention cohort. A-D**. Elastic Net coefficient paths for the four outer cross-validation folds. Each line shows the coefficient of one biomarker, or covariate across increasing regularization strength (alpha). Only features retained in the final model for the corresponding fold are shown. For each fold, the best model is represented by the dashed green line. **E**. Coefficients in the best models (one per outer fold) for each feature appearing in at least two of the best models for the intervention cohort. Red crosses indicate the coefficient estimated in each fold, and *n* indicates selection frequency across the four models ($n$ = 4 denotes selection in all four models). Numbers associated with individual coefficients indicate the within-fold rank of each feature according to resistance to penalization, with higher resistance indicating persistence of the feature coefficient under stronger regularization.

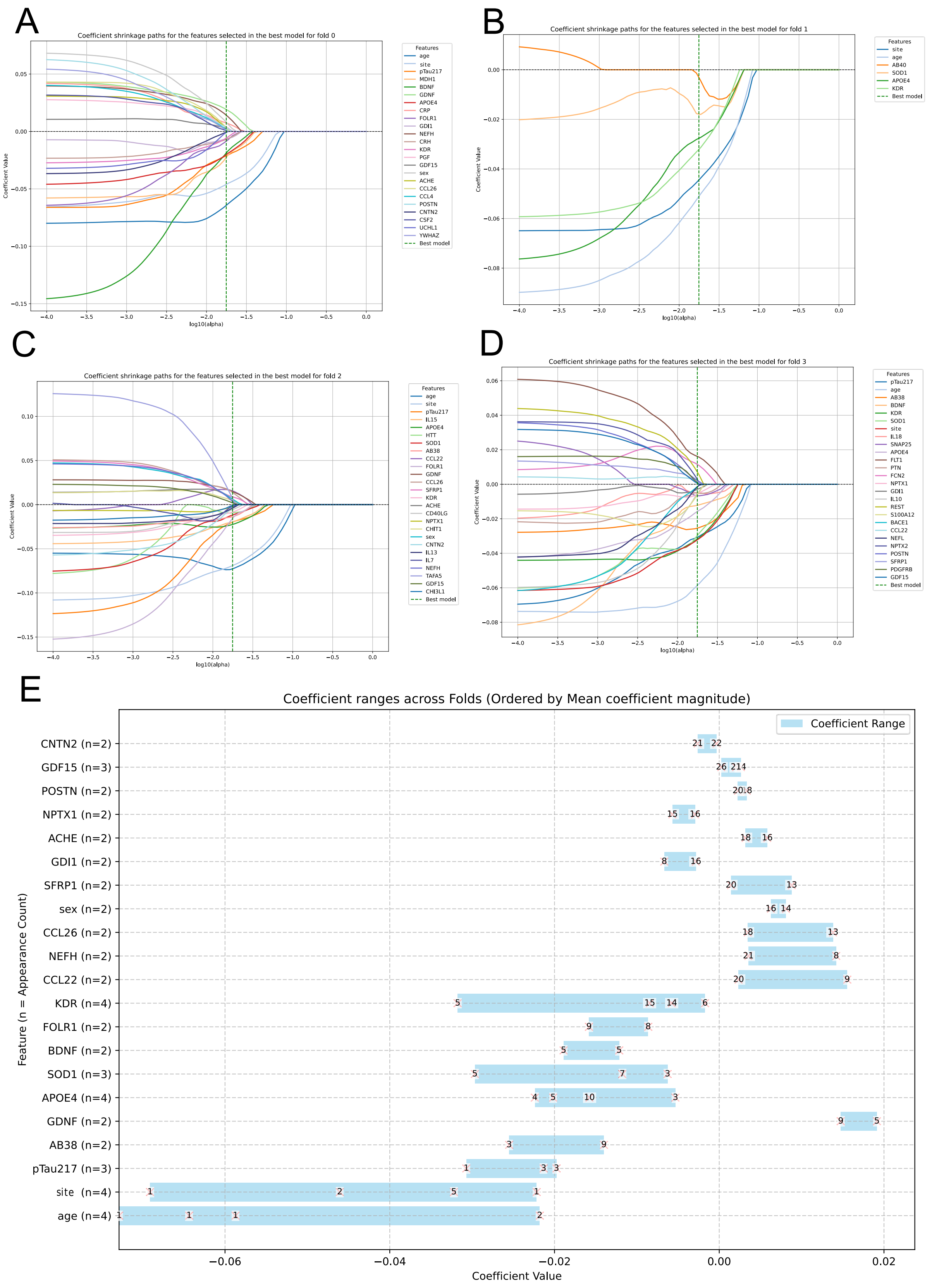

A
Coefficient shrinkage paths for the features selected in the best model for fold 0
Coefficient Value
log10(alpha)
Features
age
site
pTau217
MDH1
BDNF
GDNF
APOE4
CRP
FOLR1
GDI1
NEFH
CRH
KDR
PGF
GDF15
sex
ACHE
CCL26
CCL4
POSTN
CNTN2
CSF2
UCHL1
YWHAZ
Best model
B
Coefficient shrinkage paths for the features selected in the best model for fold 1
Coefficient Value
log10(alpha)
Features
site
age
AB40
SOD1
APOE4
KDR
Best model
C
Coefficient shrinkage paths for the features selected in the best model for fold 2
Coefficient Value
log10(alpha)
Features
age
site
pTau217
IL15
APOE4
HTT
SOD1
AB38
CCL22
FOLR1
GDNF
CCL26
SFRP1
KDR
ACHE
CD40LG
NPTX1
CHIT1
sex
CNTN2
IL13
IL7
NEFH
TAFA5
GDF15
CHI3L1
Best model
D
Coefficient shrinkage paths for the features selected in the best model for fold 3
Coefficient Value
log10(alpha)
Features
pTau217
age
AB38
BDNF
KDR
SOD1
site
IL18
SNAP25
APOE4
FLT1
PTN
FCN2
NPTX1
GDI1
IL10
REST
S100A12
BACE1
CCL22
NEFL
NPTX2
POSTN
SFRP1
PDGFRB
GDF15
Best model
E
Coefficient ranges across Folds (Ordered by Mean coefficient magnitude)
Coefficient Range
CNTN2 (n=2)
GDF15 (n=3)
POSTN (n=2)
NPTX1 (n=2)
ACHE (n=2)
GDI1 (n=2)
SFRP1 (n=2)
sex (n=2)
CCL26 (n=2)
NEFH (n=2)
CCL22 (n=2)
KDR (n=4)
FOLR1 (n=2)
BDNF (n=2)
SOD1 (n=3)
APOE4 (n=4)
GDNF (n=2)
AB38 (n=2)
pTau217 (n=3)
site (n=4)
age (n=4)
Feature (n = Appearance Count)
Coefficient Value
−0.06
−0.04
−0.02
0.00
0.02

**Supplementary Figure 5. Convergence of mixed-effects and machine-learning analyses for biomarkers associated with intervention-related cognitive response.** A. 3D surface plot of the 3-way linear mixed effect models for the biomarkers showing convergence between the mixed-effects and machine-learning analyses. The surface is produced using average value of the cofactors (or the reference when categorial). Pink: Intervention cohort. Grey: Control cohort. B. Model-predicted cognitive trajectories according to baseline biomarker level and intervention group. For visualization, low and high biomarker levels were defined using the first (Q1) and fourth (Q4) quartiles, respectively. Light grey, control group with low baseline biomarker levels; dark grey, control group with high baseline biomarker levels; light pink, intervention group with low baseline biomarker levels; purple, intervention group with high baseline biomarker levels. The whiskers correspond to the 95% confidence intervals.

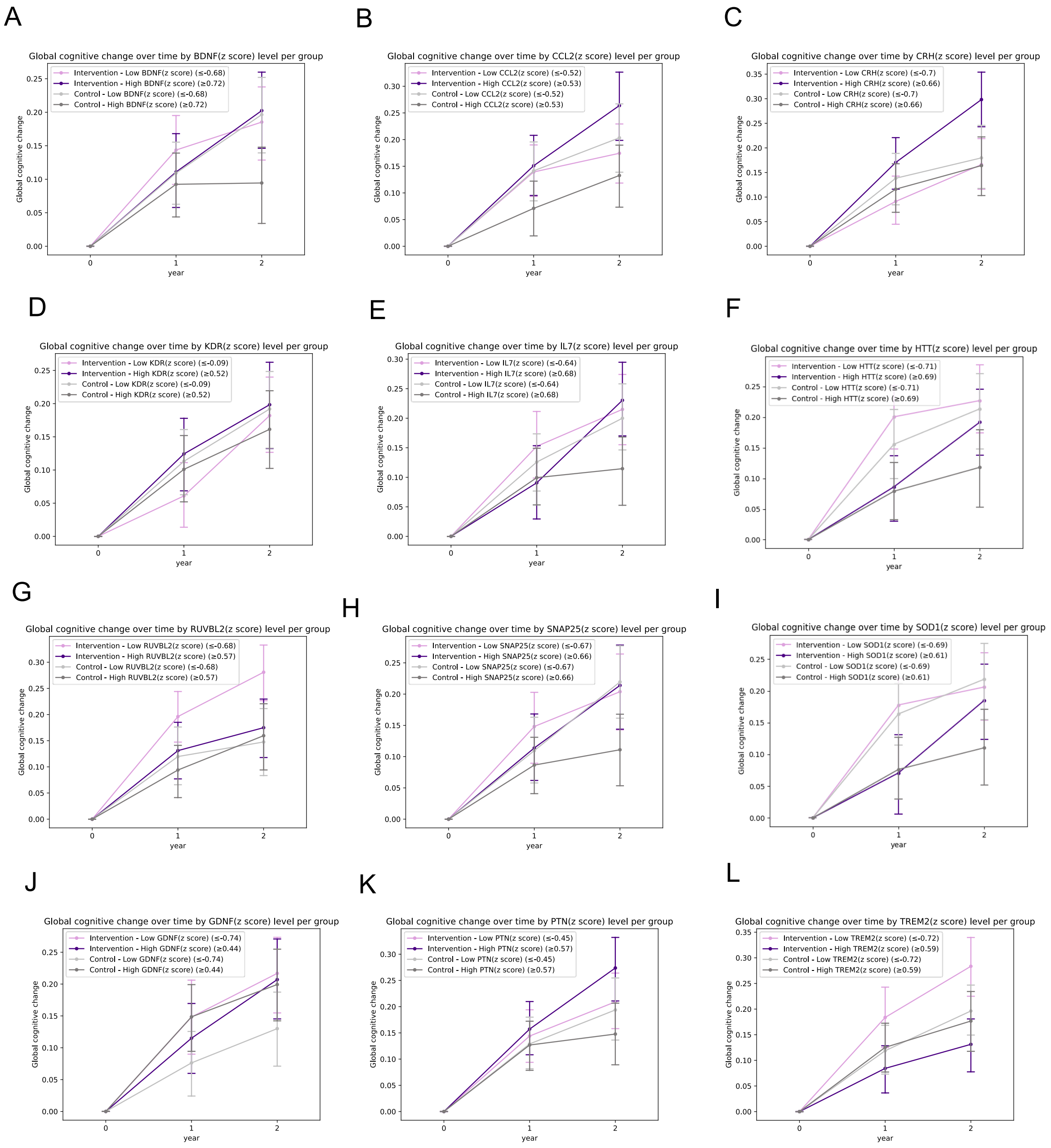


**Supplementary Figure 6. Cross-sectional correlations between plasma biomarkers.** Heatmap of the Spearman correlation matrix between baseline exploratory NULISA proteins and core blood-based ADRD biomarkers. Color intensity represents the Spearman correlation coefficient (ρ), ranging from −1 to 1. Asterisks indicate statistical significance: *$FDR\ P < 0.05$, **$FDR\ P < 0.01$ and $FDR$ ***$P < 0.001$.

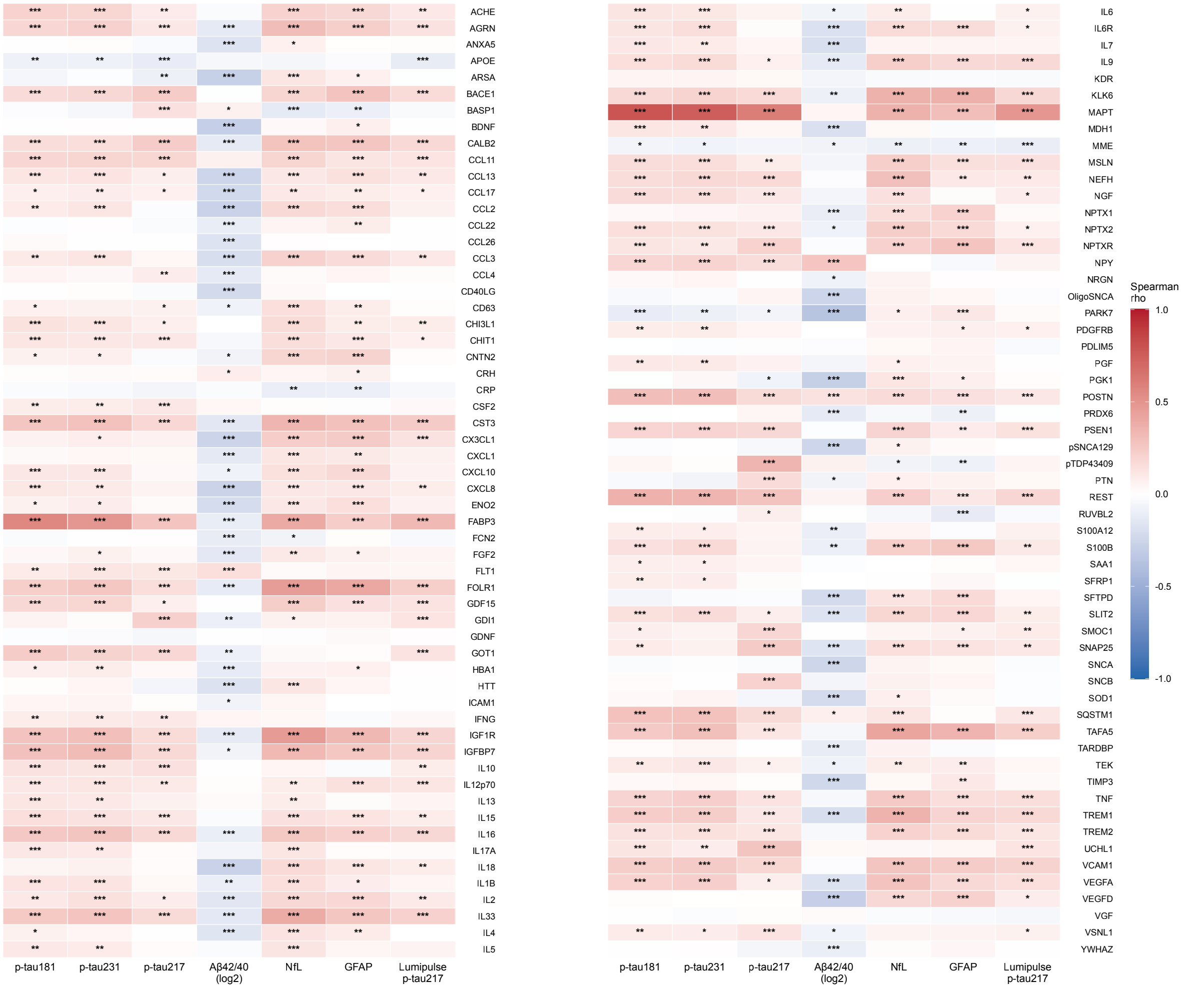
ACHE
AGRN
ANXA5
APOE
ARSA
BACE1
BASP1
BDNF
CALB2
CCL11
CCL13
CCL17
CCL2
CCL22
CCL26
CCL3
CCL4
CD40LG
CD63
CHI3L1
CHIT1
CNTN2
CRH
CRP
CSF2
CST3
CX3CL1
CXCL1
CXCL10
CXCL8
ENO2
FABP3
FCN2
FGF2
FLT1
FOLR1
GDF15
GDI1
GDNF
GOT1
HBA1
HTT
ICAM1
IFNG
IGF1R
IGFBP7
IL10
IL12p70
IL13
IL15
IL16
IL17A
IL18
IL1B
IL2
IL33
IL4
IL5
p-tau181
p-tau231
p-tau217
Aβ42/40 (log2)
NfL
GFAP
Lumipulse p-tau217
IL6
IL6R
IL7
IL9
KDR
KLK6
MAPT
MDH1
MME
MSLN
NEFH
NGF
NPTX1
NPTX2
NPTXR
NPY
NRGN
OligoSNCA
PARK7
PDGFRB
PDLIM5
PGF
PGK1
POSTN
PRDX6
PSEN1
pSNCA129
pTDP43409
PTN
REST
RUVBL2
S100A12
S100B
SAA1
SFRP1
SFTPD
SLIT2
SMOC1
SNAP25
SNCA
SNCB
SOD1
SQSTM1
TAFA5
TARDBP
TEK
TIMP3
TNF
TREM1
TREM2
UCHL1
VCAM1
VEGFA
VEGFD
VGF
VSNL1
YWHAZ
p-tau181
p-tau231
p-tau217
Aβ42/40 (log2)
NfL
GFAP
Lumipulse p-tau217
Spearman rho
1.0
0.5
0.0
-0.5
-1.0

**Supplementary Figure 7. Consort diagram.**

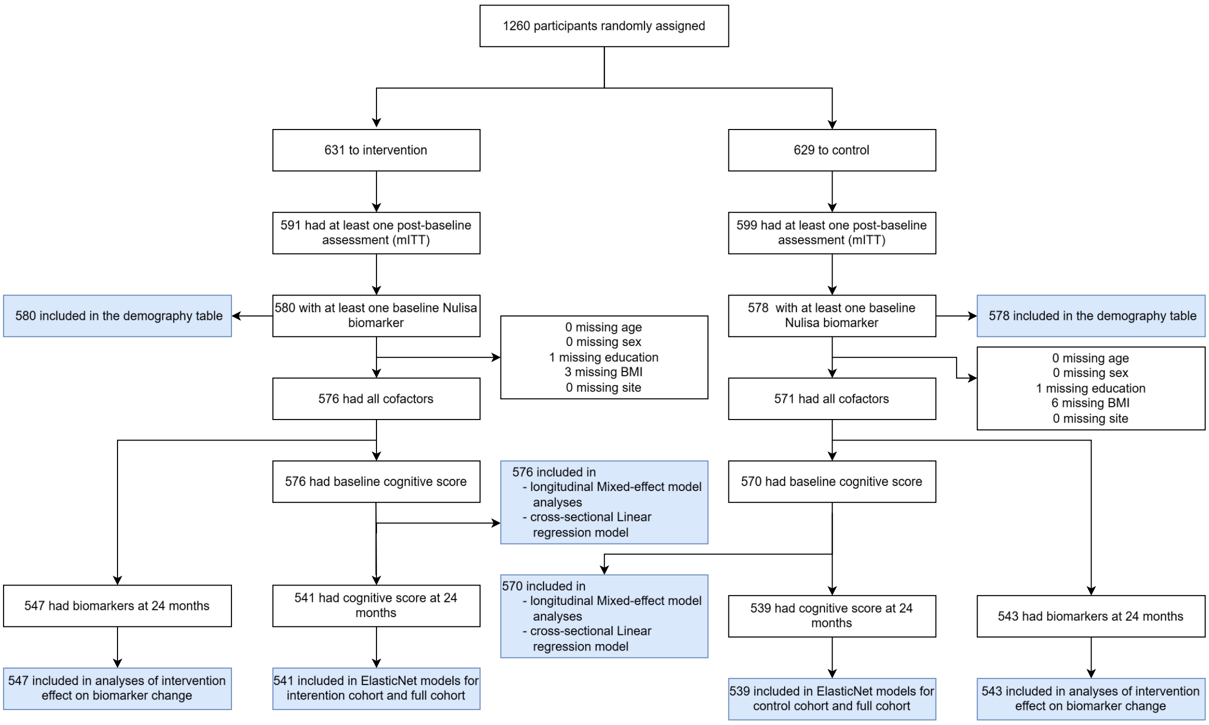
