## Supplementary material - Elastic net for "Plasma proteomics reveals signatures of cognitive aging and response to the FINGER multidomain lifestyle intervention"

### Elastic Net Validation of Biomarkers Predicting Intervention-Associated Cognitive Change

#### Supplementary Results

In addition to the mixed models, we applied Elastic Net regression as a final analytical step to identify predictive biomarkers of intervention-related cognitive response. The machine-learning models included the established blood-based ADRD biomarkers alongside the broader NULISA biomarker panel (Supplementary Table 1), as candidate predictors together with clinical and demographic variables, allowing their joint and relative contributions to prediction of cognitive change to be evaluated within a multivariable framework. Given the high correlation among biomarkers, the separate models also provide a sensitivity assessment of the stability of biomarker selection across cohorts Concordance between biomarkers identified by the two complementary approaches (Elastic Net and mixed-effect models) was used to assess the robustness of our findings and reduce the likelihood of model-specific associations. Model development and evaluation were performed using four-fold nested cross-validation, generating four independently evaluated models per cohort (intervention, control, and full cohort with biomarker x intervention group interaction), corresponding to the four outer validation folds. The selected hyperparameters, number of retained features and performance of each model are presented in Supplementary Table 8. Coefficients for all models and cohorts are presented in Supplementary Table 9.

Nineteen to twenty features were selected in models with interaction terms. The evolution of the coefficients for each of the selected features are presented in Supplementary Figure 2 A-D, while feature selection, coefficient statistics and ranking are shown in Supplementary Table 10 and Supplementary Figure 2 E. NULISA ptau217, $CRH_{group}$ and SOD1 were consistently kept across all four models while six features were selected in three of the models, including $PTN_{group}$, $CCL2_{group}$ and APOE4. Nine features were kept in half of the models. Among the retained baseline biomarkers, ptau217, ARSA, APOE4, SOD1 and CD40LG had the strongest negative coefficients in the models while the $CRH_{group}$ and $CCL2_{group}$ interactions showed highest positive coefficients. Baseline biomarkers that were most resistant to regularization were ptau217, ARSA, APOE4, SOD1 and $CRH_{group}$.

In the control cohort, APOE4 and KDR were consistently kept in all four models, while NULISA ptau217, IL15, GDF15, SOD1 were retained in three of the models. Nine features, including ACHE, GDNF, BDNF and AB38 were kept in half of the models (Supplementary table 12 and Supplementary Figure 3). Biomarkers with the strongest negative coefficients in the models were ptau217, AB38, KDR,

SOD1, APOE4 and BDNF and those with the highest positive coefficients were GDNF and CCL22. Baseline biomarkers that resisted the most to regularization were ptau217, AB38, GDNF and APOE4.In the intervention cohort, eight features were kept in all four models (age, site, CRH, CCL2, TREM2, KDR, RUVBL2, PTN) and seven in three of the models, including ACHE, GFAP, ptau231 and ptau217. Fourteen features, including sex, IGFBP7, ARSA and NEFL were retained in half of the models (Supplementary table 11 and Supplementary Figure 4). Baseline biomarkers with the strongest negative coefficients in the models were ACHE, TREM2 and IGFBP7, and those with the highest positive coefficients were CRH, CCL2, KDR and PTN. Finally, the biomarkers shown to be the most resistant to regularization were CRH, IGFBP7, TREM2, ACHE and CCL2.

Convergence between the mixed-effects and machine-learning analyses was particularly evident for CRH, CCL2 and PTN. These proteins were among the strongest positive predictors of cognitive change in the intervention-group Elastic Net models, with limited predictive contribution in the control-group models. Consistently, the three-way mixed-effects models showed more favorable associations between higher baseline CRH, CCL2 and PTN levels and longitudinal cognitive trajectories in the intervention group relative to controls (Suppl. Fig. 5). Concordant patterns across the two approaches were also observed for KDR, BDNF, GDNF, RUVBL2 and TREM2. BDNF and GDNF were preferentially retained in the control group Elastic Net models, consistent with their relatively stronger associations with cognitive trajectories in controls in the mixed-effects analyses. In contrast, RUVBL2 and TREM2 were preferentially retained in intervention group models, consistent with their intervention-dependent associations in the mixed-effects analyses. KDR was kept in both intervention and control Elastic Net models, while the mixed-effects analysis identified a significant intervention-dependent difference in its association with cognitive change.

SNAP25 and IL7 showed significant intervention-dependent associations in the mixed-effects analyses but were not consistently selected by Elastic Net. Both were highly correlated with selected predictors (SNAP25 with PTN (r=0.47) and IL7 with BDNF (r=0.79)), which may have reduced their additional contribution within the multivariable penalized models.
